# Grounding Health AI: Architecture and Evaluation of a Domain-Expert Metabolic Health Agent

**DOI:** 10.64898/2026.08.11.26359946

**Authors:** Alon Diament, Gal Sapir, Maria Gorodetski, Adva Wolf, Anna Rice, Dana Azouri, Anat Etzion-Fuchs, Dikla Gelbard Solodkin, Yeela Talmor-Barkan, Guy Lutsker, Eran Segal, Hagai Rossman

## Abstract

General-purpose language models generate fluent health reports that can fabricate derived clinical metrics. In an illustrative comparison on identical two-week CGM and meal data, leading foundation models produced reports with invented MAGE values, inflated meal counts, and unreferenced complication-risk projections — failures invisible to non-expert readers and plausible enough to mislead clinicians. We describe the HPP Personal Health Agent (PHA), a metabolic health agent that grounds generation in four layers: the Human Phenotype Project (HPP), a deep-phenotyped cohort of 13,000+ participants supplying population references and trained predictive models; 21 domain-expert tools and trained-model wrappers that compute clinical metrics and risk predictions; declarative behavioural skills that constrain what the model may claim; and 21 automated evals across 8 categories developed via a test-driven cycle in which each eval encodes a failure mode discovered during iterative development. In a 210-report matrix (14 participants × 3 prompts × 5 system conditions), the gains are largest on the system’s primary use case — meal-grounded metabolic reports, the report it was designed for — where the full system raises a deterministic form/provenance score from 0.37 (the same foundation model with no tools or skills) to 0.91; this score measures structural completeness, numerical accuracy, tool grounding, and clinical-language compliance — a necessary condition for trustworthy health reporting, with clinical quality as a complementary axis examined qualitatively. A skills-vs-tools decomposition shows the two layers act on different axes: tools drive numerical accuracy (≈14% → 90% of reported metrics correct), while the declarative skills add most of the remaining gain in citations, completeness, and structure (tools alone recover only part of the gap, 0.49 from the same 0.37 baseline). The lift generalises beyond the primary use case — to a second metabolic prompt (0.72) and a cardiovascular extension (0.70), each from a 0.37–0.39 baseline. The architecture extends across clinical domains: adding a SCORE2 cardiovascular risk tool and a corresponding skill — with no changes to orchestration, eval harness, or existing tools — produced a cardiovascular risk report from the same system. Trustworthy domain-specialised health AI is a systems design problem: deep-phenotyped cohort data, domain-expert tools and models, and eval-driven development together form a replicable pattern.

## 1. Introduction

Large language models can now produce fluent, clinically plausible health reports from raw patient data. Given a two-week continuous glucose monitor (CGM) trace, a diet log, and basic demographics, a frontier LLM will return a well-structured document complete with glycaemic metrics, risk assessments, and dietary recommendations. The problem is that much of it may be wrong. When we provided identical input data to two leading foundation models (Claude Sonnet 4.5 and GPT-5.2), both generated reports that read convincingly but diverged sharply from ground truth on derived clinical quantities. One model reported a mean amplitude of glycaemic excursions (MAGE) of 41.0 mg/dL — “within the acceptable range” — when the validated value was 84.8 mg/dL, roughly double. The other claimed to have analysed 172 meals when the input contained 70. Both offered specific complication-risk projections with no model, no uncertainty, and no methodology (Section 6.3). These are not edge cases; they are the expected behaviour of systems that lack the computational and data infrastructure to ground their claims (Figure 1).

**Figure 1.**
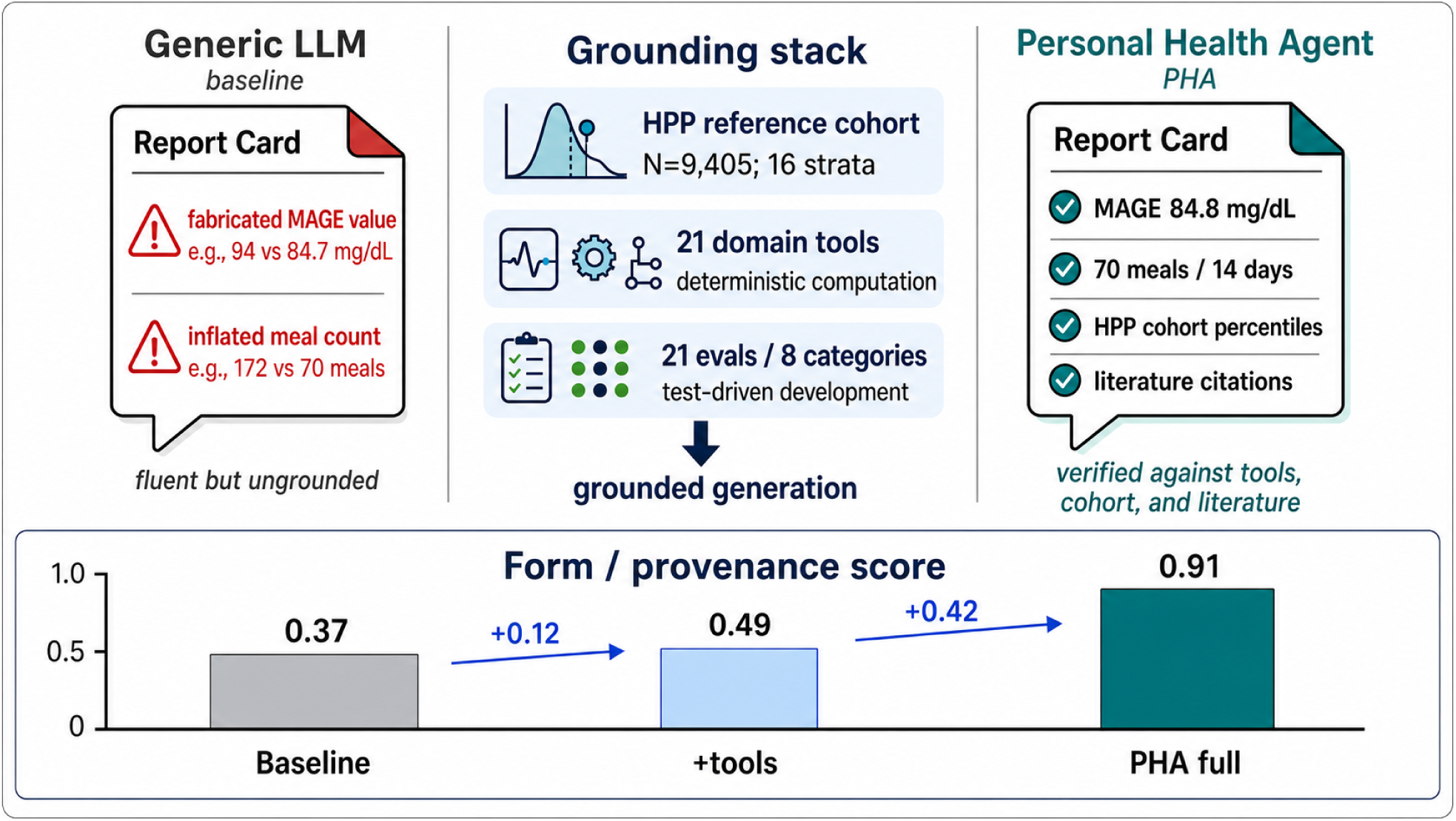
| Grounded versus ungrounded health reporting. *Left:* a generic LLM baseline produces a fluent but fabricated report — MAGE estimated at 94 mg/dL (validated: 84.8) and 172 meals claimed (actual: 70). *Centre:* the grounding stack — HPP reference cohort (N = 9,405; 16 strata), 21 domain-expert tools, and 21 evals across 8 categories developed via a test-driven cycle. *Right:* the PHA report, with all values verified against tools, cohort, and literature. *Bottom:* mean deterministic form/provenance score across the 14-participant matrix — baseline 0.37, adding tools alone 0.49, full system 0.91.

This failure mode — confident fabrication of health-relevant quantities — is particularly dangerous because non-expert users cannot reliably detect it. In a large-scale evaluation of a wearable-data health agent, (Merrill et al. 2026) found that while the system achieved 84% numerical accuracy, users struggled to distinguish grounded responses from plausible-sounding but incorrect ones. (Heydari et al. 2025) reported a similar asymmetry: clinical experts identified quality differences with high inter-rater agreement, but non-expert participants could not. The implication is clear: fluency is not a proxy for accuracy, and the burden of correctness cannot rest on the end user.

Recent work addresses parts of this gap from two directions — tool augmentation for clinical computation and benchmarks for post-hoc model assessment (Section 2) — but these threads remain largely separate, and the existing frameworks are designed for post-hoc measurement rather than development-time iteration.

Our position is that trustworthy domain-specialised health AI is an integration problem. Trustworthiness requires four co-designed layers — a deep-phenotyped reference cohort, a suite of domain-expert tools and models, declarative behavioural skills that constrain what the LLM may claim, and an evaluation harness that drives development rather than assesses it after the fact — each closing a failure mode the others cannot. We instantiate this architecture as the HPP Personal Health Agent (PHA), built on the Human Phenotype Project (HPP) — a deep-phenotyped cohort of over 13,000 participants with clinical-grade CGM, dietary logging, blood tests, body composition, and multi-omic measurements. The system comprises a comparatively simple architecture: one ReAct orchestrator, 21 domain-expert tools and trained-model wrappers, and declarative behavioural skills expressed as markdown files — coupled with a test-driven evaluation framework of 21 automated evals across 8 categories that drove iterative development rather than assessed it post hoc.

We make four contributions. First, we demonstrate that deep-phenotyped cohort data — population reference distributions, trained predictive models, and multi-modality cohort matching from the HPP — enables personalised health insights that no general-purpose model can replicate, and we show how to make this data accessible to an LLM through 21 domain-expert tools and declarative behavioural skills. Second, we provide ablation and baseline evidence quantifying what each layer contributes: removing skills degrades groundedness and clinical language compliance; removing literature grounding eliminates evidence-based citations; and stripping the entire tool suite produces confident fabrication of clinical quantities. Third, we describe a practical eval-driven development methodology, including a taxonomy of domain-specific failure modes — from silent tool failures that produce plausible but meaningless predictions, to skill examples that inadvertently teach the LLM to hallucinate. Fourth, the architecture is portable: extending PHA from metabolic to cardiovascular risk required one new tool and one new skill, with no changes to orchestration, evals, or existing tools — an initial demonstration of a pattern in which additional clinical domains can be integrated by adding domain-specific content while reusing the same underlying infrastructure.

## 2. Related Work

Recent work on LLM-based personal health agents has demonstrated increasing sophistication in both architecture and evaluation. PHIA (Merrill et al. 2026) showed that ReAct agents with generic tools (code interpreter, web search) can answer open-ended questions over wearable data, achieving 84% numerical accuracy in a large-scale human evaluation. PH-LLM (Khasentino et al. 2025) took a complementary fine-tuning approach, specialising Gemini on sleep and fitness time-series to exceed human expert performance on domain benchmarks — demonstrating domain-specific gains but at the cost of portability across health domains. Health-LLM (Kim et al. 2024) established baseline capabilities for wearable health prediction across 12 LLMs and 10 tasks, finding that context enhancement improves performance substantially but that even augmented models struggle with derived clinical metrics. Most recently, (Heydari et al. 2025) presented the most comprehensive such system to date: a multi-agent Gemini 2.0 system with three specialist sub-agents evaluated across 10 benchmark tasks with over 7,000 expert annotations. Evaluation in these systems was conducted after the system was complete — often rigorously, but as a measurement tool rather than a development driver.

Separately, a growing body of work has focused on tool augmentation and evaluation infrastructure for medical LLM agents. AgentMD (Jin et al. 2024) demonstrated that equipping LLMs with over 2,000 clinical calculators improved risk prediction accuracy from 40.9% to 87.7%, and (Goodell et al. 2025) found that task-specific clinical tools produced 5.5–13× error reductions compared to generic code interpreters — together establishing that domain-specific tools, not just general-purpose capabilities, are essential for reliable clinical computation. On the evaluation side, MedAgentBench (Jiang et al. 2025) introduced a FHIR-compliant interactive environment with 300 patient-specific tasks; HealthBench (Arora et al. 2025) scaled to 5,000 multi-turn conversations evaluated against 48,562 physician-written rubric criteria; and MedHELM (Bedi et al. 2025) proposed a clinician-validated taxonomy spanning 121 tasks across 22 subcategories. These benchmarks provide valuable measurement of model capabilities but are designed for post-hoc assessment rather than development-time iteration.

Our work integrates these elements into a single system: a comparatively simple architecture — one ReAct orchestrator with 21 domain-expert tools and trained-model wrappers and declarative behavioural skills — coupled with a TDD-like evaluation framework of 21 automated evals that drove development iteratively rather than assessed it after the fact. Critically, the entire system is grounded in the Human Phenotype Project (Reicher et al. 2025), a deep-phenotyped cohort of 13,000+ participants whose population reference data enables personalised benchmarking that no general-purpose model can replicate. We provide not only the agent and its tools but also the evaluation methodology, ablation evidence for each architectural layer, and empirical lessons on failure modes — aiming to offer a replicable pattern for building trustworthy domain-specialised health AI.

## Methods

### 3. The Human Phenotype Project as Data and Models Platform

The system described in this paper is built on top of the Human Phenotype Project (HPP), a longitudinal deep-phenotyping study of over 13,000 participants conducted at the Weizmann Institute of Science. We briefly describe the cohort and the derived data products that the agent relies on; full details are provided in (Reicher et al. 2025).

#### 3.1 Cohort and Phenotyping Depth

HPP participants undergo clinical-grade measurements spanning continuous glucose monitoring (14-day CGM via FreeStyle Libre), prospective dietary logging (timestamped food items with nutritional composition), sleep monitoring and wearable devices, blood tests (complete blood count, metabolic panel, HbA1c, lipid profile), body composition (DXA), blood pressure, anthropometrics, and multi-omic profiling (gut microbiome metagenomics, serum metabolomics via Nightingale, untargeted metabolomics, proteomics via Olink, and low-pass whole-genome sequencing). A subset of participants return for longitudinal follow-up visits, enabling prospective outcome tracking — the 2-year dysglycaemia risk model is trained on confirmed glycaemic outcomes at the 2-year follow-up to predict future dysglycaemia from baseline CGM features (Supplementary S1). This combination of multi-modal depth, clinical-grade measurement quality, and longitudinal design distinguishes the HPP from publicly available wearable datasets, which typically offer one or two modalities at consumer-grade resolution.

#### 3.2 Population Reference Data

Two derived data products enable personalised benchmarking — the central capability that separates this system from general-purpose health LLMs.

##### Age– and sex-stratified percentile references

For 46 CGM-derived metrics, we pre-computed empirical percentile distributions (5th, 10th, 25th, 50th, 75th, 90th, 95th) across 9,405 HPP participants stratified into 16 strata (eight 5-year age bins from 30–70 × two sexes) (Keshet et al. 2023). These enable the population percentile tool (Supplementary S1) to report, for example, that a participant’s mean glucose of 157 mg/dL places them at the 100th percentile among 298 age– and sex-matched males — a statement that requires real cohort data and cannot be approximated from textbook values.

##### K-Nearest Participants (KNP)

A privacy-preserving cohort matcher identifies the most similar participants across seven data modalities (CGM, anthropometrics, dietary patterns, body composition, blood pressure, clinical labs, and demographics). The matching algorithm operates on normalised feature vectors and returns aggregate statistics of the matched cohort without exposing individual records. This enables multi-dimensional population context: rather than comparing a participant’s glucose in isolation, the system can characterise the metabolic profile of similar individuals across modalities. KNP comparisons ground population context in real cohort data, enabling multi-dimensional comparisons of the kind the baseline models attempted to generate without a reference cohort (Section 6.3).

#### 3.3 Trained Predictive Models

The agent integrates five predictive tools, four trained on HPP data and one implementing a published clinical algorithm: current dysglycaemia classification, prospective 2-year dysglycaemia risk, metabolic age, visceral adipose tissue (VAT) estimation, and SCORE2 cardiovascular risk. The HPP-trained Random Forest models report performance metrics and SHAP-based feature attributions alongside their predictions; the prospective dysglycaemia model also uses conformal prediction to communicate uncertainty. The suite further includes GluFormer (Lutsker et al. 2026) for short-horizon glucose forecasting and a postprandial glucose response (PPGR) predictor for meal optimisation. Model cards, training sizes, validation metrics, and implementation details are provided in Supplementary S1.

## 4. System Design

### 4.1 Architecture Overview

The system follows a three-stage pipeline (Figure 2): orchestration, analysis, and deterministic rendering. A **ReActOrchestrator** (a DSPy ReAct agent) iteratively selects and executes tools, adapting its strategy based on observations — retrying on errors, skipping unavailable analyses, or adjusting parameters. Each tool call returns lightweight metadata (10–15 curated scalars) to the orchestrator for reasoning, while heavy data objects (DataFrames, figures) are persisted to an artifact store and referenced by ID. A **ReActAnalyzer** then synthesises the accumulated tool outputs into a coherent report narrative, querying artifacts through bounded accessor tools that enforce a strict information budget — the analyzer never receives a full dataset, only bounded views. The analyzer produces the complete report content; a deterministic **Reporter** module then combines this narrative with CGM visualisations into a formatted PDF, without further LLM involvement. Behavioural skills (Section 4.3) are injected at both the orchestrator and analyzer stages.

**Figure 2.**
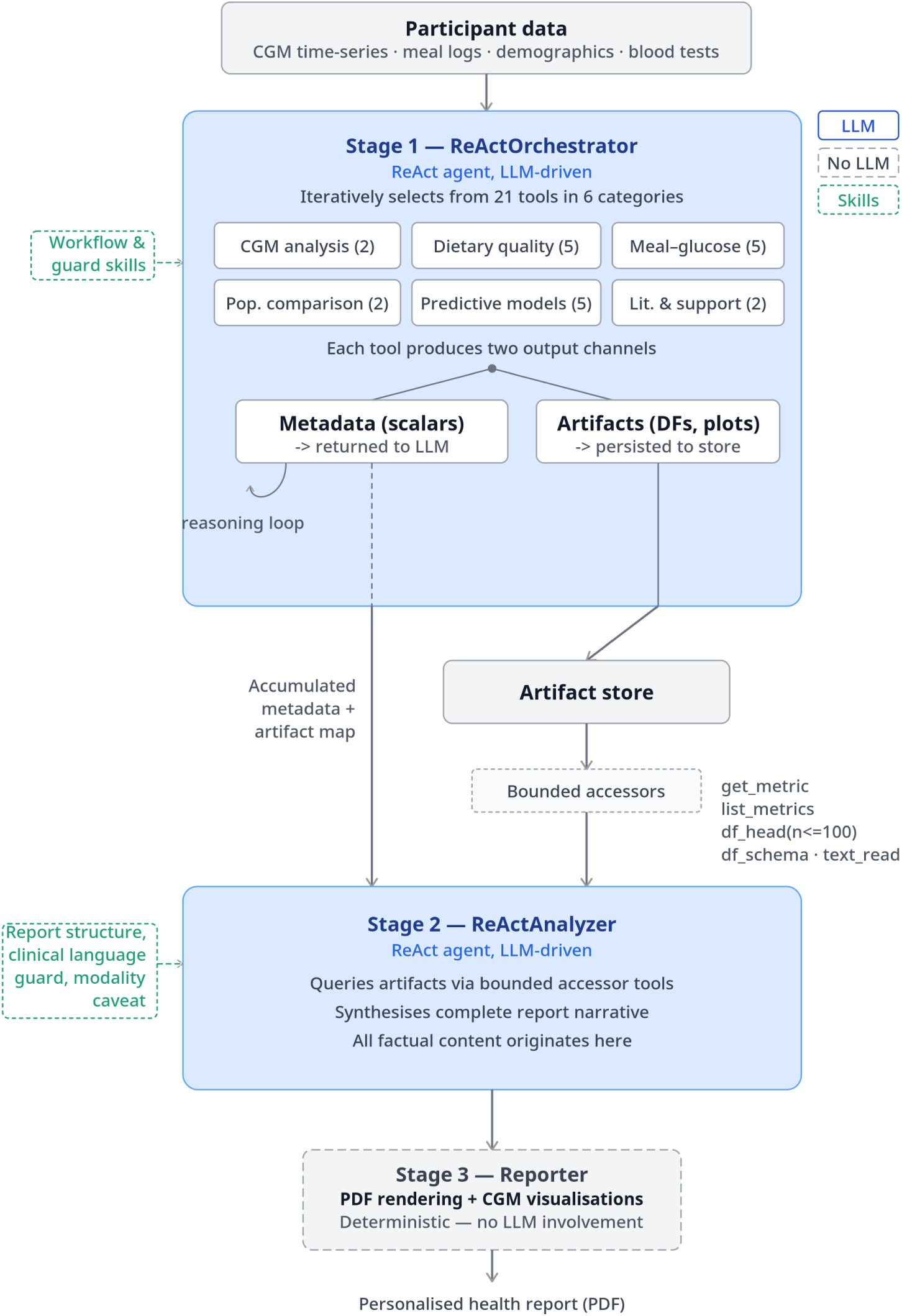
| System architecture. Three-stage pipeline — ReActOrchestrator (21 domain tools in 6 categories; CGM analysis 2, dietary quality 5, meal-glucose response 5, population comparison 2, predictive models 5, literature & support 2; two output channels: metadata to the LLM, artifacts to a store), ReActAnalyzer (bounded artifact accessors, produces all factual report content), and a deterministic Reporter (PDF + CGM visualisations, no LLM). Behavioural skills are injected at the orchestrator and analyzer stages.

### 4.2 Domain-Expert Tool Suite

The health agent operates through a curated suite of 21 domain-expert tools (tools and trained-model wrappers), each encapsulating a well-defined analytical or predictive capability. Rather than routing all analysis through a single LLM call, the system decomposes health data analysis into discrete, composable operations — from raw data ingestion and quality control through clinical metric computation, population-level comparison, and predictive modelling. This modular architecture was motivated by three considerations. First, each tool implements validated, domain-specific logic (e.g., consensus CGM metrics computed via the iglu library (Broll et al. 2021) (Python port: iglu-python), the SCORE2 cardiovascular risk algorithm (SCORE2 working group and ESC Cardiovascular risk collaboration et al. 2021)) that can be independently tested and updated without affecting the rest of the system. Second, the LLM orchestrator (Section 4.1) selects and chains tools dynamically based on the clinical question and available data, enabling flexible, patient-specific workflows without hard-coded pipelines. Third, each tool exposes a standardised interface — typed inputs, structured metadata output, and file-based artifact references — allowing independently authored tools to compose without coupling (Supplementary S1). Behavioural skills (Section 4.3) further constrain how the LLM interprets and reports tool outputs, enforcing clinical language standards and grounding requirements.

The 21 domain tools are organised into six functional categories (full per-tool contracts in Supplementary S1):

- **CGM analysis** (2): parsing, consensus metrics (Broll et al. 2021) — turns raw CGM streams into computed quantities rather than LLM-estimated summaries
- **Dietary quality** (5): meal ingestion, QC, FCDB enrichment, dietary indices (Zhan et al. 2024), narrative generation — separates food items from meals and prevents unsupported nutrition claims
- **Meal-glucose response** (5): meal window construction, CGM alignment, PPGR quantification, short-horizon forecasting (GluFormer; CGM × diet model), dietary optimisation — connects diet to measurable glycaemic response instead of free-text dietary inference
- **Population comparison** (2): age/sex-adjusted HPP percentile ranking, matched-cohort summaries — grounds comparisons in real cohort data with disclosed sample sizes
- **Predictive models** (5): dysglycaemia classification, 2-year risk, metabolic age, VAT estimation, CVD risk — replaces unsupported projection with transparent model-backed prediction (training details in Supplementary S1)
- **Literature & support** (2): multimodal data loading, medical literature retrieval — preserves data provenance and citation support for clinical interpretation

The 21-tool count excludes reporting/visualisation helpers, which package presentation artefacts rather than domain measurements.

A distinguishing feature of this tool suite is the integration of rule-based deterministic methods with LLM-augmented reasoning within several tools. For example, the diet quality control tool applies 16 rule-based validators (completeness, Atwater consistency, range checks) and supplements them with LLM-based semantic judgement for ambiguous cases. Similarly, the meal segmentation tool uses deterministic time-gap thresholds to identify meal boundaries and delegates borderline cases to an LLM assessor that evaluates temporal, compositional, and physiological plausibility. This hybrid approach preserves reproducibility for well-defined cases while leveraging LLM flexibility for edge cases that resist simple heuristics.

Detailed tool interface specifications (structured output envelopes, artifact ID conventions, and framework adapters), diet quality control validation rules (16 deterministic + 2 LLM validators), and the temporal meal segmentation algorithm are provided in Supplementary S1.

The three representative tools highlighted in the supplement — SCORE2, population_percentile, and the 2-year dysglycaemia risk model — illustrate the suite’s three domain contributions: a published clinical algorithm wrapped behind the standard tool interface, population-reference computation against the HPP cohort, and a longitudinal HPP-trained predictive model. The remaining tools follow the same interface.

### 4.3 Skills as Behavioural Constraints

Tools provide the system with computational capabilities; skills constrain how the language model uses and communicates the results. A skill is a markdown file — no code, no API calls — that is injected into the system prompt at either the orchestrator stage, the analyzer stage, or both. The system uses 12 skills in three categories: **guard skills** that enforce invariants (e.g., clinical language-guard maps forbidden diagnostic terms to data-grounded alternatives; modality-caveat-guard requires caveats when mentioning unavailable data), **report-structure skills** that define the output format and clinical voice, and **workflow skills** that guide tool selection and multi-tool coordination for specific analytical domains.

One important finding from our development process was that skills must be calibrated to tool capabilities. For each claim type a skill encourages, there must be a tool that produces structured data to support it. We identified three outcomes when auditing skill–tool alignment: (a) *grounded* — a tool computes the relevant metric; (b) *groundable* — the data exists in an artifact but the skill does not direct the LLM to retrieve it; and (c) *ungroundable* — no tool computes the quantity, so the LLM will hallucinate it. The third category proved especially dangerous when skill examples contained illustrative data. A skill example showing time-of-day glucose patterns — a metric that no tool in the suite computes — taught the LLM to fabricate matching patterns in its output. Removing ungroundable examples eliminated this failure mode without any changes to the tool suite or orchestration logic (Section 7).

The meal_cgm_response skill illustrates the grounded category in practice. Its YAML header declares the injection points and the exact tools whose outputs the skill is allowed to reference; its FORBIDDEN block then enumerates the hallucinations the skill prohibits, forcing every claim back to tool metadata. The key invariant is that item counts from meal_parser must not be reported as meal counts. That instruction is the textual trace of a real failure: an earlier version of the system reported the raw item count as the meal count, a failure analogous to the baseline-LLM inflation reported in Section 1. The prohibition is small, specific, and entirely expressible in markdown; no code change was required. The full skill is reproduced in Supplementary S2.

## 5. Evaluation Framework

Before writing evals, we specified the tasks they would test: the expected input modalities, required tool chains, and success criteria for each proposed capability. Rather than evaluating the system only after it was built, we adopted a test-driven development (TDD) approach in which each automated eval encodes a failure mode discovered during iterative development. The cycle — *identify failure* → *write eval* → *fix tool or skill* → *confirm no regressions across existing evals* — drove the system’s evolution (Figure 3). Because each eval captures a failure we had already observed and corrected, a high pass rate reflects how thoroughly we closed known failures rather than correctness on unseen cases; the meaningful evidence is therefore the comparison across conditions (Section 6.2) and the ground-truth comparisons below, which check reported values against independently validated references.

**Figure 3.**
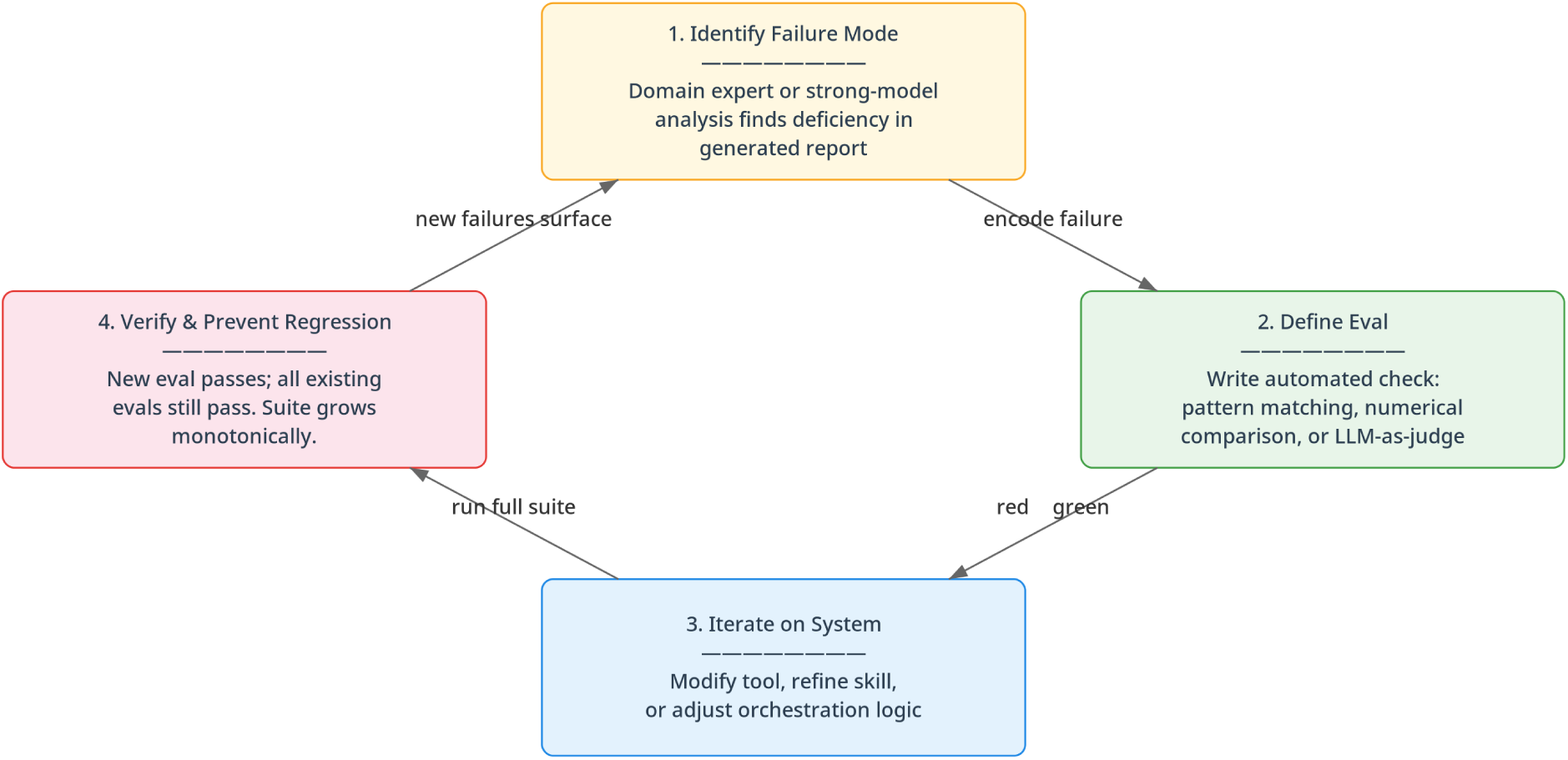
| Eval-driven (test-driven) development cycle. Each automated eval encodes a failure mode discovered during development: a domain expert or strong model identifies a deficiency, the failure is encoded as an automated check (pattern matching, numerical comparison, or LLM-as-judge), the tool or skill is fixed, and the full suite is re-run so the eval suite grows monotonically.

We evaluate structured personalised health reports that exercise the full tool suite (CGM analysis, dietary assessment, population comparison, predictive models, and literature grounding). The main experiment (Section 6.1) generates these reports from three realistic prompts over a fixed participant set, providing a controlled, reproducible evaluation surface — same participant data, the same prompts across conditions, observable improvement over iterations — while still constraining outputs to a single report format. The evals use three complementary methods: (1) **deterministic** pattern matching for structural properties (required sections, forbidden terms, cohort N disclosure); (2) **ground-truth comparison** for computational accuracy (e.g., reported mean glucose within 1 mg/dL of the validated value); and (3) **LLM-as-judge** assessment for properties that resist deterministic checking (appropriate hedging, narrative coherence, grounded recommendations). Task stratification, expert annotation workflow, related-eval framing, and the full eval registry are provided in Supplementary S3.

### 5.1 Eval Categories

The 21 evals are organised into 8 categories, each targeting a cluster of failure modes discovered during error analysis. Table 1 summarises each category with its motivating failure, representative evals, and eval type.

**Table 1.** Eval categories and representative failure modes.

| Category | Motivating Failure | Representative Evals | Type |
| --- | --- | --- | --- |
| 1. Groundedness | Percentiles reported without cohort size; temporal claims unverifiable against CGM trace | Temporal anchor check for glucose event claims; cohort N disclosure; modality caveats | Det. + LLM |
| 2. Clinical language | “CRITICAL,” “SEVERE HYPOGLYCEMIA” applied to healthy participants’ sensor artefacts | Forbidden term scan; hedging verification for interpretive claims | Det. + LLM |
| 3. Completeness | Critical CGM metrics or entire sections omitted when tools returned partial results | Required metrics presence (mean, GMI, CV, TIR); section completeness | Det. |
| 4. Meal/PPGR | Meal-glucose associations claimed without diet data; PPGR hallucinated without tool output | Tool usage verification; coverage disclosure (meals analysed, %); hallucination detection vs. tool output | Det. + GT + LLM |
| 5. Predictions | Predictions reported without uncertainty; SHAP omitted; values fabricated after silent tool failure | Tool invocation; value accuracy vs. tool output; uncertainty reporting; SHAP presence | Det. + GT + LLM |
| 6. Citations | “Evidence-based” labels without actual references; guideline bodies named without specific criteria | Citation presence for interpretive claims; citation–claim alignment | Det. + LLM |
| 7. Negative patterns | System invented clinical problems for healthy participants; alarming language for routine findings | No problem invention (healthy profiles); no alarming language; appropriate reassurance | Det. + LLM |
| 8. Population comparison | Cohort comparisons without sample size, matching criteria, or small-N caveats | Cohort N disclosure; matching criteria reported; small cohort caveat (N < 20) | Det. + LLM |

These evals measure what we term *form*: structural compliance, numerical accuracy, tool usage, and linguistic constraints. We acknowledge that *essence* — clinical appropriateness, narrative coherence, and utility to the patient — resists automated evaluation and ultimately requires expert review and user studies (Section 8).

## 6. Results

We report the systematic evaluation as evidence on *form* and *provenance* — the structural, numerical, tool-grounding, and linguistic properties the eval suite measures (Section 5). These are a lower bound on the trustworthiness that motivates the architecture, not a demonstration of clinical quality; clinical *essence* is addressed only by the qualitative case study in Section 6.3 and is left to expert review (Section 8).

### 6.1 Setup

The main experiment is a matrix of 14 participant samples × 3 prompts × 5 system conditions (210 reports), all generated with Claude Opus 4.6. Samples are HPP participants with ground-truth CGM and demographic data; 11 carry prospective diet logs and 3 do not, the latter serving as modality-caveat stress tests under the diet-dependent prompt. The three prompts are realistic first-person user requests rather than internal specifications: a general metabolic scorecard (metabolic_scorecard), a diet-and-CGM action request (meal_grounded_action), and a cardiovascular-risk scorecard (cardiovascular_risk_scorecard).

The meal-grounded action prompt is the system’s primary use case: the metabolic report the tool suite and skills were designed around. The metabolic scorecard and the cardiovascular-risk prompt test generalisation — to a second metabolic intent and to a new clinical domain, respectively. We therefore report the meal-grounded result as the main result (Section 6.2) and treat the other two prompts as generalisation.

The five conditions isolate the contribution of skills against tool availability; the two ablations (No-lit-skill, No-meal-skill) are skill-guidance ablations, not tool-denial ablations — the full tool suite is reachable in every condition:

- **Baseline** — the same Opus 4.6 foundation model given the prompt and the raw record, with no tools and no skills.
- **Full** — the full tool suite plus all skill prompts.
- **Tools-only** — the full tool suite available, but no skill prompts injected.
- **No-lit-skill** — all skills except the literature-context skill; the literature-search tool remains available.
- **No-meal-skill** — all skills except the six meal/diet skills; the meal/diet tools remain available.

FCDB enrichment and meal optimisation are excluded from the main matrix, meal text is summary-only, and external-provider replication is deferred.

The eval registry exposes 21 evals — 18 deterministic checks (pattern-matching and ground-truth comparison, both run without an LLM) and 3 LLM judges; every report is run against the full registry. The headline form/provenance scores (Section 6.2, Figures 4 and S2) aggregate the deterministic checks only; the three LLM judges are reported separately and treated as provisional (Section 6.2). Two scoring conventions keep conditions comparable. First, tool-sensitive evals are split into a **content score** (does the report make the claim?) and a **tool-execution score** (was the claim backed by a tool call recorded in the trace?). Second, conditions are not penalised on dimensions whose tool they cannot use: baselines receive no tool-execution penalty, and the no-lit and no-meal ablations are skipped on their removed-tool dimensions.

**Figure 4.**
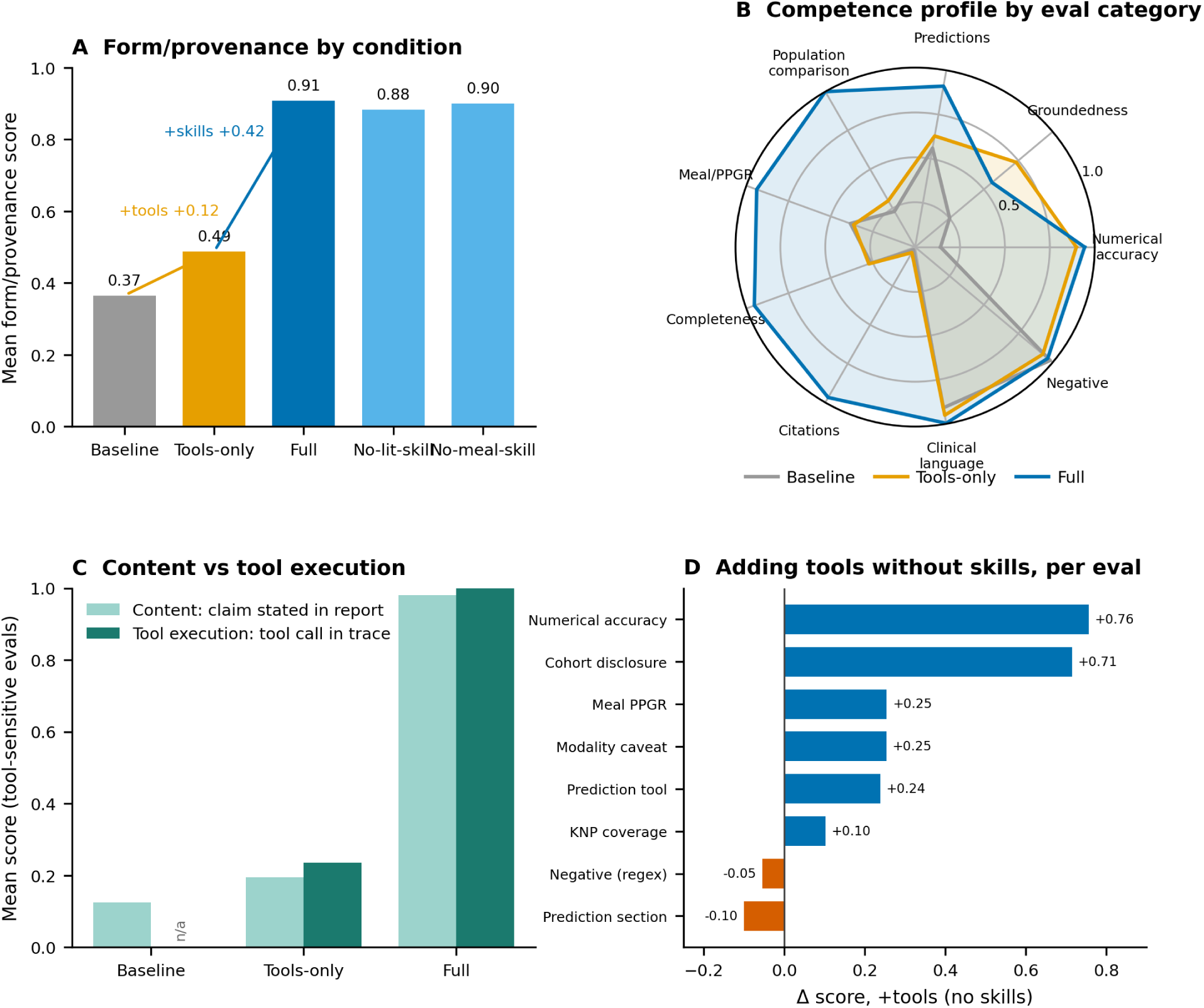
| Form/provenance evaluation on the primary use case (meal-grounded reports; 14 participants, Opus 4.6). **(A)** Mean form/provenance score by condition; the full system reaches 0.91 from a 0.37 baseline, and the decomposition shows that adding the tool suite without skills recovers little (+0.12) while the skills supply the bulk (+0.42). **(B)** Competence profile across the nine eval axes — the eight categories of Table 1 plus numerical accuracy, shown separately — for baseline, Tools-only, and Full; the baseline polygon collapses on numerical accuracy, citations, population comparison, completeness, and meal/PPGR; numerical accuracy is restored by tools alone, the rest require skills, and clinical-language and negative-pattern compliance are already high without the system. **(C)** Content (does the report state the claim?) versus tool execution (is the corresponding tool call present in the trace?), averaged over the five tool-sensitive evals; the baseline has no tools (n/a) and tool execution rises only once skills are present. **(D)** Per-eval effect of adding tools without skills (Δ score, baseline → no-skills; the 8 of 16 evals with |Δ| ≥ 0.05 are shown, the remainder change by less than 0.05): tools alone sharply raise the accuracy and grounding evals (numerical accuracy +0.76, cohort disclosure +0.71) but regress a couple of form evals (prediction-section −0.10, negative-regex −0.05). All quantities are deterministic form/provenance measures, not clinical-quality judgments.

The matrix baseline differs from the deep-dive evidence in Section 6.3: here the baseline is the same Opus 4.6 model stripped of tools and skills, whereas Section 6.3 is an illustrative single-participant comparison that also uses baseline report artifacts and an external-model benchmark deliverable.

The 14 participants (7F/7M, mean age 58 years [range 32–72], mean BMI 27 kg/m²) were selected to span the full glycaemic range: six have normoglycaemic CGM profiles (GMI < 5.7%), five fall in the pre-diabetic to early diabetic range (GMI 5.7–6.7%), and three have established uncontrolled glycaemia (GMI ≥ 7.0%). Two participants in the mid-range show HbA1c ≥ 7.1% despite near-normoglycaemic CGM, consistent with medically managed diabetes. Eleven carry prospective food logs; three do not, serving as modality-caveat stress tests under the diet-dependent prompt.

### 6.2 Results: Primary Use Case (Meal-Grounded Reports)

On meal-grounded reports the full system raises the deterministic form/provenance score from 0.37 — the same foundation model with no tools or skills — to 0.91 (Figure 4A). Removing *all* skill guidance collapses it to 0.49. Removing a single skill family moves it only slightly: No-lit-skill (0.88) mainly drops the literature citations from the report, and No-meal-skill (0.90) remains high because the meal *tools* still run without the meal skills — so a single-family ablation chiefly changes what is reported, not whether the computation happens.

#### Skill guidance, not tool availability, drives form/provenance compliance

Tools without any skill prompts (Tools-only, 0.49) recover only a small part of the 0.37 → 0.91 gap (+0.12 of the +0.54 total); the skills supply the rest. The two layers act on different axes (Figure 4B). Tools make grounded computation *possible*: numerical accuracy is almost entirely a tool effect, rising +0.76 (0.14 → 0.90) when tools are added and flat thereafter, so reported CGM metrics are roughly 14% accurate without tools and 90% with them, with or without skills. The declarative skills then determine whether the report *states* it: at the skill step, citations rise 0.93 points, population comparison +0.70, completeness +0.68, meal reporting +0.57. Clinical-language and negative-pattern compliance are already high at baseline (clinical language ≈0.91, negative patterns ≈0.98), so the skills’ distinctive contribution here is citations, structure, and disclosure rather than more careful wording — latest SOTA LLMs avoid explicitly alarmist phrasing by default, and the deterministic checks target specific forbidden terms the model naturally omits. The more substantive failures documented in §6.3 (fabricated diagnoses, unsupported risk grades) use hedged language that evades term-matching; those are not what these checks measure.

Conditions are averaged over slightly different eval sets, since an eval whose inputs are absent is skipped rather than scored zero (15–17 of the 17 deterministic evals, depending on condition). On the 13 evals scored in every condition × participant cell the gap narrows from +0.54 to +0.48 and the tool contribution to performance grows from +0.12 to +0.16, leaving the ordering and the skills-dominate conclusion unchanged.

#### Content versus tool execution

Splitting the tool-sensitive evals into the claim made versus the tool call behind it (Figure 4C) shows the grounding directly. The full system both makes these claims and backs almost all of them with recorded tool calls (content 0.98, tool execution 1.0). The baseline scores low on both (it has no tools to call), and, notably, tools-without-skills makes little use of the tools it has (content 0.20, tool execution 0.23): the skills are what trigger the tool calls and surface their outputs.

#### Adding tools without skills is not uniformly beneficial (Figure 4D)

The accuracy and grounding evals rise sharply at the tool step (numerical accuracy +0.76, cohort disclosure +0.71, prediction-tool +0.24), but a couple of form evals *regress* — prediction-section −0.10, negative-regex −0.05 — recovering only once skills are reinstated. Capability without behavioural guidance lets the agent assert more and structure less. The same compute-versus-report division appears within categories: invoking the predictor is tool-driven while reporting it with uncertainty and SHAP is skill-driven, and the digital-twin/KNP comparison is essentially skill-gated.

#### LLM judges (provisional)

The three LLM judges (all using Claude Sonnet 4.5 as the judge model) are held out of the form/provenance scores above and analysed on their own. The citation judge separates the conditions sharply and skill-dependently (≈0.01 at baseline, 0.00 without skills, 1.0 with the full system), whereas the groundedness and negative-pattern judges separate only weakly and rate the baseline highly (groundedness ≈0.78 and negative-pattern ≈0.91 at baseline, against ≈0.95–0.97 for the full system). That leniency sits in tension with the concrete baseline fabrications in Section 6.3, and is the reason the judges are excluded from the headline scores; we treat them as provisional pending a judge-versus-human agreement check on an annotated subset, and base no form/provenance claim on them.

##### Generalisation across prompts

The pattern holds beyond the primary use case (Figure S2). On a general metabolic scorecard and on a cardiovascular-risk prompt the full system reaches 0.72 and 0.70 respectively, against baselines of 0.37–0.39 — the baseline-to-full lift is present on all three prompts and largest on the meal-grounded use case the system was built for. Across all five conditions (Figure S2) the skill ablations track the full condition closely; the one exception — No-meal-skill edging above the Full condition on the metabolic scorecard — is an artefact of that condition being scored over fewer (meal-specific) evals, not a real gain. The cardiovascular prompt is a domain extension (Section 7) and the weakest of the three; we read it as evidence of portability, not of cardiovascular clinical quality.

### 6.3 Baseline Comparison: General-Purpose LLM vs. Full System

This section is an illustrative case study, not part of the systematic matrix (Sections 6.1–6.2). It examines Participant A because the ground truth, PHA report, same-prompt baseline report, early baseline artifact, and external-model benchmark deliverable are all inspectable (Supplementary S5).

To isolate the contribution of the agent architecture — tools, skills, cohort data, and structured computation — we compared baseline reports (the same foundation model run on the raw record without tools or skills; the Baseline condition from §6.1) with the PHA report for the same patient record: 13 days of CGM readings, 70 logged meals with nutritional composition, and demographic data (age, sex, BMI, HbA1c). The baseline reports received the record but lacked the agent layer: no CGM analysis library, no cohort database, no meal-glucose alignment pipeline, no predictive models, and no curated domain skills. This setup tests a specific hypothesis: that access to the raw data, combined with a frontier LLM’s medical knowledge, is insufficient for reliable health reporting.

The baseline reports were fluent, well structured, and clinically plausible on first reading. The failures emerged in quantities that require computation, provenance, or refusal when inputs are missing. Table 2 summarises the divergences across the five dimensions examined below.

**Table 2.**
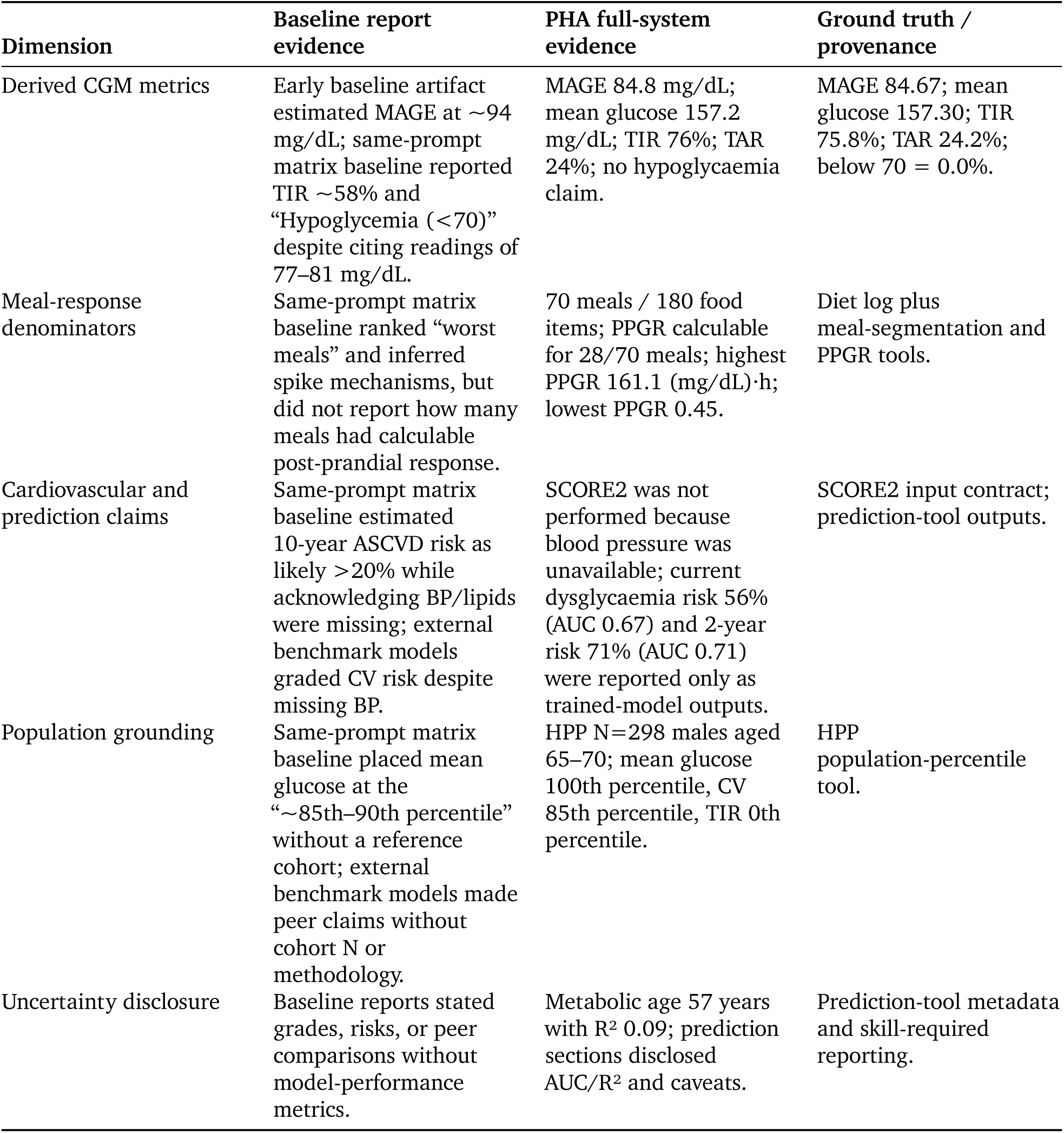
Baseline reports versus PHA on the illustrative single-participant case (Section 6.3). PHA values are rounded as reported; ground-truth values from validated tool outputs.

#### Computed metrics and denominators

The early baseline artifact reported a fabricated MAGE estimate of approximately 94 mg/dL, while the computed value is 84.67 mg/dL (Service et al. 1970). The same-prompt matrix baseline showed a larger error on time-in-range, reporting approximately 58% rather than 75.8%, and labelled “Hypoglycemia (<70)” as rare while pointing to readings of 77–81 mg/dL, which are not hypoglycaemic. PHA reported the rounded computed values and separated meal claims from meal-alignment coverage: 70 logged meals, 180 food items, and PPGR calculable for only 28 meals. The point is not that every baseline number is wrong; it is that the report gives the reader no reliable way to tell extracted quantities from computed ones or valid denominators from inferred ones.

#### Grounded refusals and risk quantification

The most important cardiovascular result is a refusal. The matrix baseline estimated 10-year ASCVD risk as likely greater than 20% despite acknowledging that exact calculation required lipids and blood pressure. In the external-model benchmark, every non-PHA model assigned a cardiovascular grade from diabetes status and diet patterns despite missing blood pressure. PHA instead stated that SCORE2 could not be performed without blood pressure and shifted to actionable next steps: obtain blood pressure and lipid data. Where PHA did make predictions, the values came from trained tools: 56% current dysglycaemia risk with AUC 0.67, 71% two-year dysglycaemia risk with AUC 0.71, and metabolic age 57 years with R² 0.09.

#### Population grounding and transparent uncertainty

The same-prompt baseline reported peer percentiles without naming a reference population. The external-model benchmark showed the same pattern: peer claims such as “worse than most men your age” or “bottom 25%” appeared without N, cohort definition, or method. PHA drew on 298 age– and sex-matched HPP males to report exact percentiles: 100th for mean glucose, 85th for CV, and 0th for TIR. It also disclosed weak model performance when appropriate, including the R² 0.09 metabolic-age caveat. This is the calibrated behaviour the eval framework is designed to enforce: compute when a tool exists, refuse when required inputs are absent, and disclose uncertainty when predictions are model-based.

Taken together, these domains illustrate a consistent pattern. The PHA architecture does not produce reports that are uniformly better than the baselines in every dimension — the baseline reports occasionally generate more fluent prose, more detailed behavioural recommendations, or qualitative observations our current tools do not compute. What the architecture provides is groundedness: every numerical claim traces to a computation, every comparison traces to real data, and every prediction comes with a performance caveat. In each domain, the absence of grounding led the baselines not to silence or expressed uncertainty, but to confident extrapolation beyond the available evidence.

We extended the comparison to four additional frontier models run without the agent layer — GPT-5.4, Claude Opus 4.6 (PHA’s own foundation model), Gemini 3.1 Pro, and Perplexity Sonar Pro — across two prompts on the same single participant; these observations come from benchmark deliverables and are not part of the scored matrix. The pattern was consistent. GPT-5.4 understated time above 140 mg/dL (approximately 50% versus 67.4%). Claude Opus 4.6 reported TIR 68% versus 75.8%, TAR 28% versus 24.2%, and described 220–270 mg/dL spikes as routine despite only 0.8% of readings exceeding 250 mg/dL. Claude Opus 4.6 also fabricated a reactive hypoglycaemia diagnosis from a single 77 mg/dL reading — below no standard threshold — and derived medication-class advice from it (insulin timing and dose review); this is qualitatively distinct from a numerical error, and a clinician acting on it could inappropriately reduce diabetes medication in a patient with zero hypoglycaemic episodes. Gemini and Perplexity made unverifiable peer claims without a cohort N, and all four external models graded cardiovascular risk despite missing blood pressure. At the same time, the external models sometimes identified useful qualitative patterns, such as dawn-phenomenon signatures or meal-stacking effects. That is a capability gap in the current tool suite, not an argument against the architecture: closing it means adding validated tools for those patterns rather than relying on ungrounded narrative inference.

## Discussion and Limitations

### 7. Discussion

The HPP Personal Health Agent demonstrates that grounded, trustworthy metabolic health reporting is achievable through a layered architecture of cohort data, domain-expert tools, behavioural skills, and eval-driven development — and that the same architecture extends to new clinical domains without structural change. Two findings from our development process merit discussion; the second speaks more broadly to system portability.

#### Skills must be calibrated to tool capabilities

Simple markdown files proved surprisingly effective at controlling LLM behaviour — the clinical-language-guard skill eliminated diagnostic terminology, and the population-comparison skill reliably triggered cohort N disclosure. But this effectiveness cuts both ways. As defined in Section 4.3, skills partition into grounded, groundable, and ungroundable categories; the third proved especially dangerous in practice. A skill example showing time-of-day glucose patterns — a metric no tool computes — taught the LLM to fabricate matching patterns. Removing ungroundable examples eliminated this hallucination without any code changes.

#### Domain content carries the system; the orchestration harness is scaffolding

Two migrations during development tested this. In the first, we replaced the agent framework (DSPy → Claude Agent SDK); all 21 domain tools, 12 skills, and 21 evals transferred without modification, and only the orchestration adapter layer was rewritten. In the second, we extended the system from metabolic health to cardiovascular risk reporting: we added one new tool (a SCORE2 (SCORE2 working group and ESC Cardiovascular risk collaboration et al. 2021) cardiovascular risk calculator) and one new skill (a cardiovascular report template), with no changes to orchestration, existing tools, existing skills, or the evaluation harness. The resulting report is not a separate pipeline that silos cardiovascular reasoning: the same CGM analysis, cohort-matching, and predictive tools built for the metabolic setting compose their outputs with the new SCORE2 calculation in a single coherent document. A representative assessment for Participant B — full report in Supplementary S6 — illustrates the integration: glucose variability (CV 10.85%, 5.5th percentile among 616 age-and sex-matched HPP males), a 2-year dysglycaemia projection (24%, AUC 0.71), metabolic age (50.3 years, R² 0.09), and 10-year CVD risk (3.56%, ESC 2021 low-risk category) appear in a single document, each with its model-performance caveat. Clinical-domain specialisation and framework-level concerns live in separate layers from the orchestration itself, so domain extension becomes a matter of authoring content rather than re-engineering the system. Investment should flow primarily into the cohort, tools, skills, and evals, since the orchestration layer is the most likely part to be replaced as the field matures.

An extension of the preceding two findings, motivated by the preliminary cross-model comparison in Section 6.3: each model we tested exhibited its own fabrication signature — quiet understatement in one, alarmist escalation in another, invented clinical-inference chains with medication-class speculation in a third. A grounded system does not need to anticipate every model’s failure mode, because the numerical and population-comparison claims do not come from the model. A pragmatic consequence is that foundation-model upgrades can be evaluated against the same eval harness and deployed without re-learning each release’s hallucination tendencies, which now affect only the narrative layer rather than the numerical one.

### 8. Limitations and Future Work

Our evaluation framework measures *form* well — citation presence, tool invocation, numerical accuracy, forbidden terminology, structural completeness. *Essence* — clinical appropriateness, narrative coherence, utility to the patient — resists automated evaluation and ultimately requires clinical expert review and prospective user studies. The 21 evals and the failure taxonomy in Section 4.3 are a lower bound on trustworthiness, not a definition of it.

The system is grounded in a single, distinctive cohort. The HPP is a deep-phenotyped Israeli population; generalisation to other demographics has not been tested, and the pre-computed percentile references and HPP-trained predictive models inherit the cohort’s composition. The current system produces a static metabolic health report — interactive coaching, the planned next system, is out of scope for this paper. We report results on a small evaluation participant set and have not conducted a prospective study with clinicians or patients, so external validity beyond the reported cases is limited. The cross-model comparison in Section 6.3 is preliminary, based on a single participant (Participant A) and two prompts across four external models; the qualitative patterns reported should be read as early observations whose specific conclusions may shift as the sample grows.

Our ablation isolates skill guidance against tool availability but includes no tools-off / skills-on condition, so the contribution of skills *without* tools is not directly measured — the skill effects in Section 6.2 are measured on top of a tool-equipped system. This is compounded by the eval surface: several groundedness, prediction, and meal evals require a tool call to pass, so a skills-only system would be structurally capped on those axes regardless of report quality. The tool-versus-skill decomposition should therefore be read as conditional on this design and sample, not as a general claim about skills in isolation.

LLM-as-judge evals have their own failure modes — sensitivity to prompt phrasing, anchoring on surface features, cost — and we triangulate wherever possible against deterministic pattern checks and ground-truth comparison. Latency is non-trivial: a complete report requires roughly 12 orchestrator iterations and 200–400 seconds end-to-end (somewhat longer on current model versions), which rules out synchronous interactive use at the present cost point. A pragmatic production architecture would combine deterministic pipelines for the stable computational core with an agent for routing and narrative synthesis; we see this hybrid path as the shortest route to deployment. Future work includes an interactive Q&A agent over the same tool suite, expansion to additional HPP data modalities, a prospective study with clinicians and participants, and a test of whether declarative skills reduce the phrasing-driven inconsistency recently documented for medical QA by (Yun et al. 2026) — skills constrain the claim surface the LLM may invoke, which should plausibly narrow such variability, but we have not tested this directly.

### 9. Conclusion

Trustworthiness in health AI is a systems design problem. Deep-phenotyped cohort data provides the foundation; domain-expert tools and models, behavioural skills, and automated evals build trustworthiness on top. The architecture is replicable: extending from metabolic to cardiovascular health required one new tool and one new skill, without changes to orchestration, evaluation, or any existing component. As foundation models improve, the grounding layer ensures that gains in language capability translate to better narratives rather than more plausible fabrications. The static report used as the evaluation substrate here is one instantiation of the architecture; the same cohort, tools, skills, and eval harness can ground health chatbots, insight generators, or other LLM-backed interfaces — the report format was chosen because its bounded structure makes grounding auditable, not because the architecture requires it. The primary investment should be in the cohort, tools, skills, and evals — the components that encode domain knowledge — since orchestration is the layer most likely to be replaced as the field matures.

## Code and Data Availability

A minimal reference implementation of the architecture is available at github.com/alondmnt/hpp-health-agent under an MIT licence. Cohort data access is governed by the Human Phenotype Project consortium; see humanphenotypeproject.org.

## Ethics

HPP participants sign informed consent on arrival at the research site. Identifying details are removed before computational analysis. The study is conducted according to the Declaration of Helsinki and was approved by the Weizmann Institute of Science IRB, approval number 2392-4.

## Data Availability

HPP cohort data are not publicly available; access is governed by the Human Phenotype Project consortium and restricted by participant consent. The data are available to research institutions on request (https://humanphenotypeproject.org/). A minimal reference implementation with a synthetic example dataset is openly available at https://github.com/alondmnt/hpp-health-agent.

## Acknowledgements

We thank the Human Phenotype Project participants and the Human Phenotype Project study team for building and maintaining the cohort infrastructure that made this work possible. E.S. is supported by the Crown Human Genome Center; the Larson Charitable Foundation New Scientist Fund; the Else Kroener Fresenius Foundation; the White Rose International Foundation; the Ben B. and Joyce E. Eisenberg Foundation; the Nissenbaum Family; Marcos Pinheiro de Andrade and Vanessa Buchheim; Lady Michelle Michels; Aliza Moussaieff; grants funded by the Minerva Foundation, with funding from the Federal German Ministry for Education and Research; the European Research Council; and the Israel Science Foundation. The funders had no role in study design, data collection and analysis, decision to publish or preparation of the manuscript.

## Author Contributions

A.D., G.S., M.G. and H.R. conceived the study, designed the analyses, developed the methodology, performed the investigation, interpreted the results and wrote the manuscript; A.W. and A.R. developed domain-expert tools, reviewed the analyses and critically revised the manuscript; D.A., A.E.F., D.G.S., Y.T.B., G.L. and E.S. reviewed the analyses, interpreted the results and critically revised the manuscript. All authors approved the final manuscript.

## Supplementary Material for: Grounding Health AI: Architecture and Evaluation of a Domain-Expert Metabolic Health Agent

### S1. Tool contract table

All tools expose the same external contract to the ReAct orchestrator: a named tool with typed inputs, bounded metadata returned to the language model, and artifact identifiers for larger objects such as tables, figures, traces, and markdown reports. Rows use the tool names exposed to the orchestrator. The 21-row table excludes reporting/visualisation helpers. Those helper tools package presentation artefacts rather than domain measurements; for example, generate_cgm_meal_visualizations is documented in the skill in Section S2 because it constrains report assembly. The wrapper and artifact envelope are generic scaffolding; the table below documents the domain contract for each tool, including the quantity computed and the provenance needed to interpret its output.

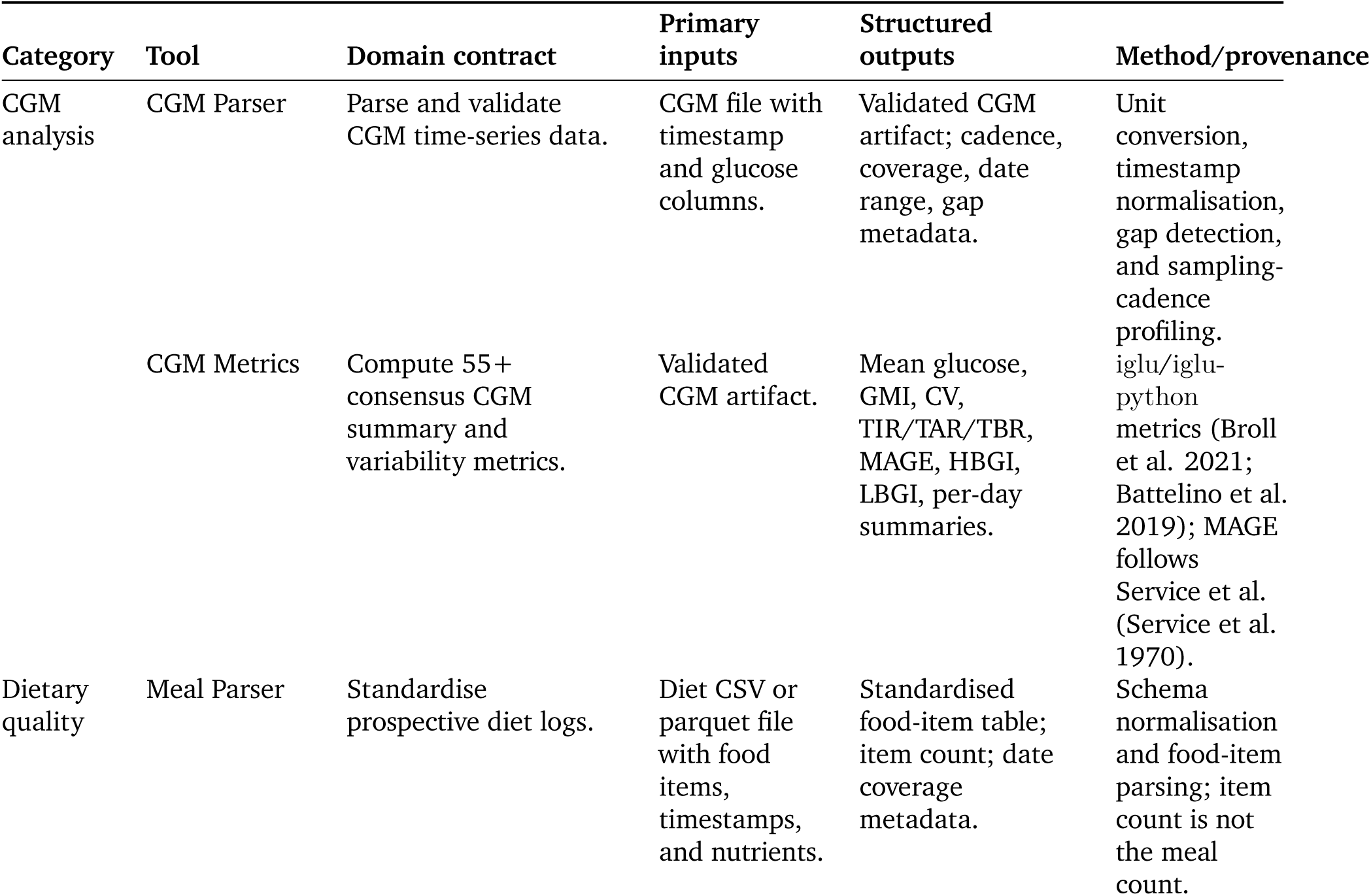
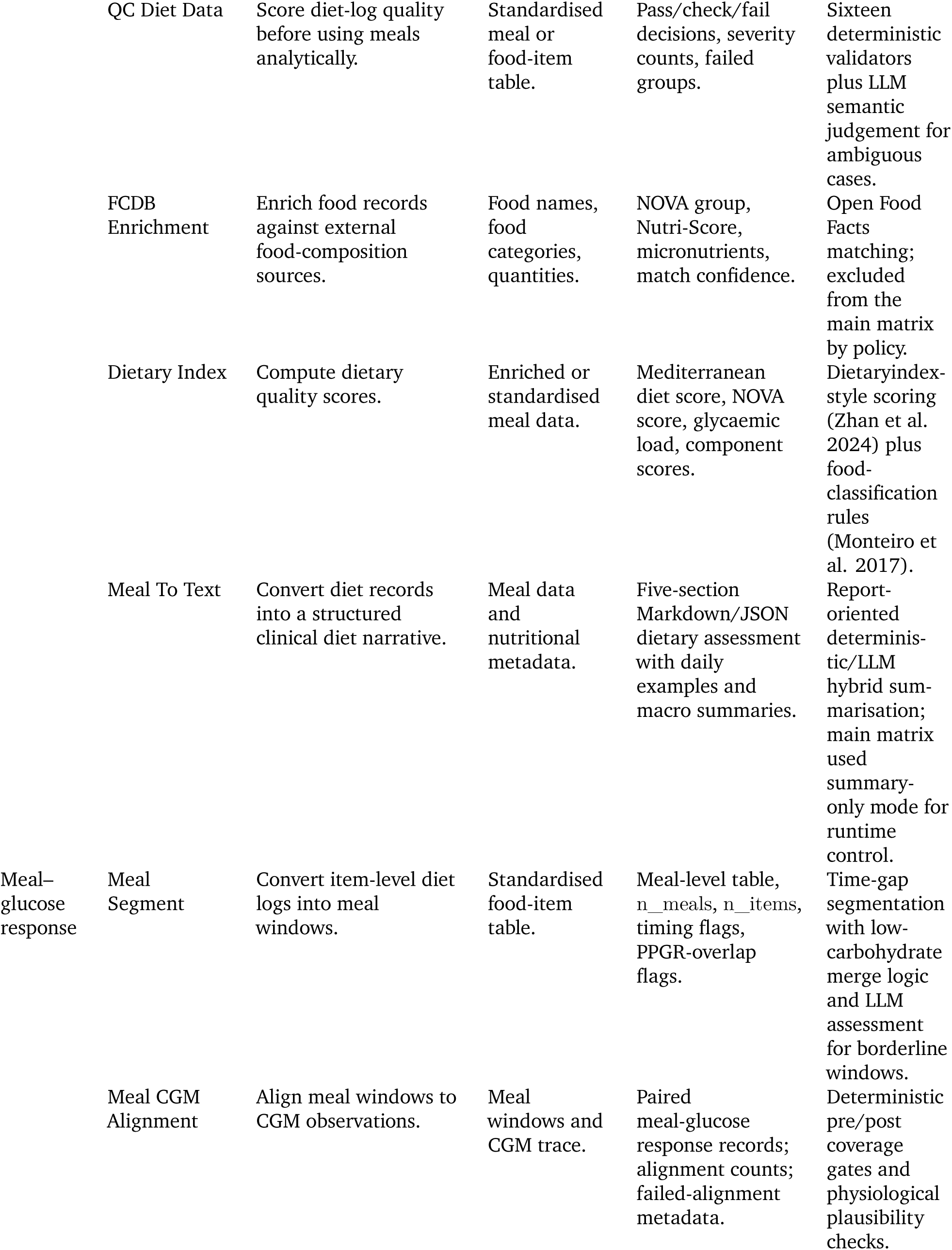
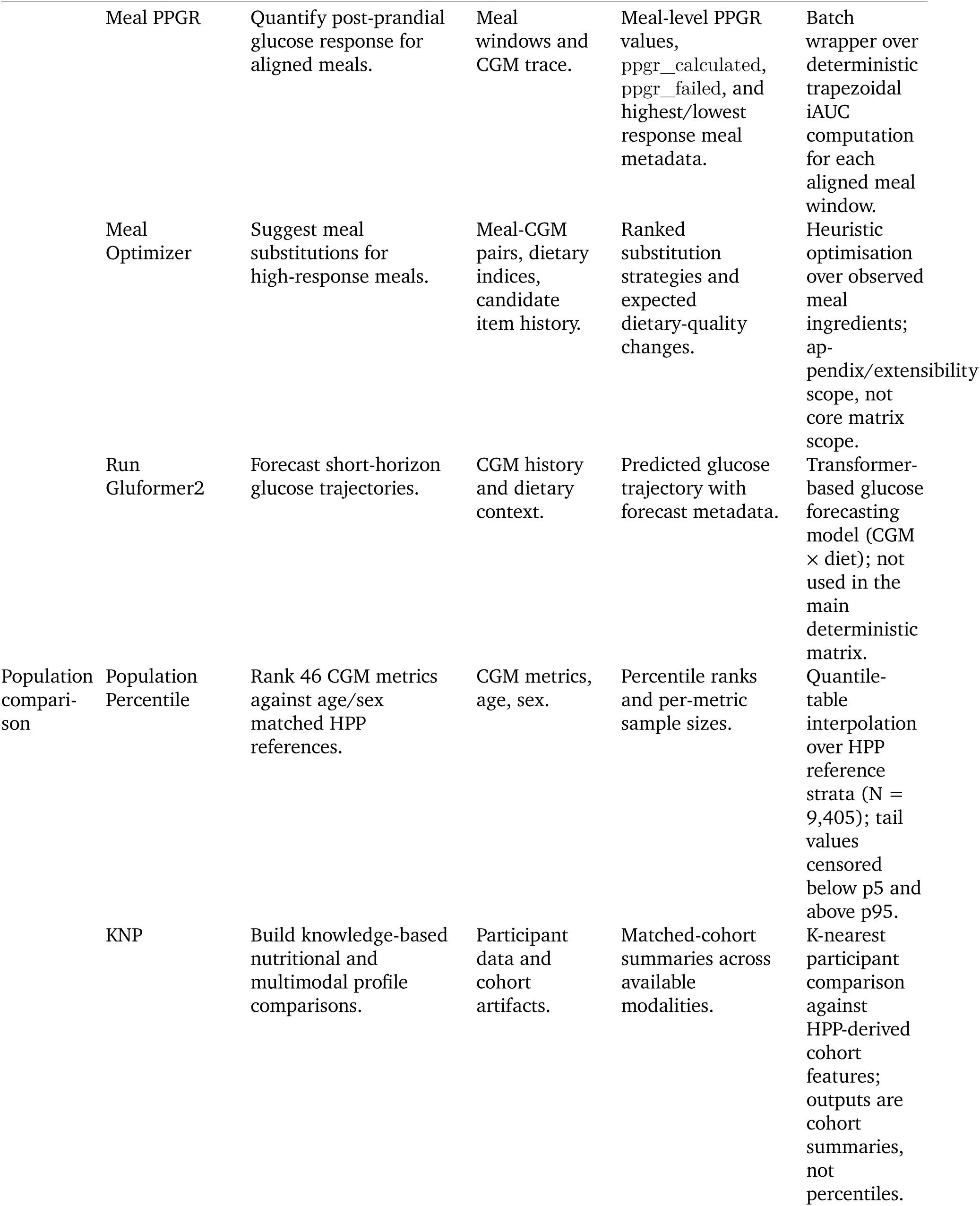
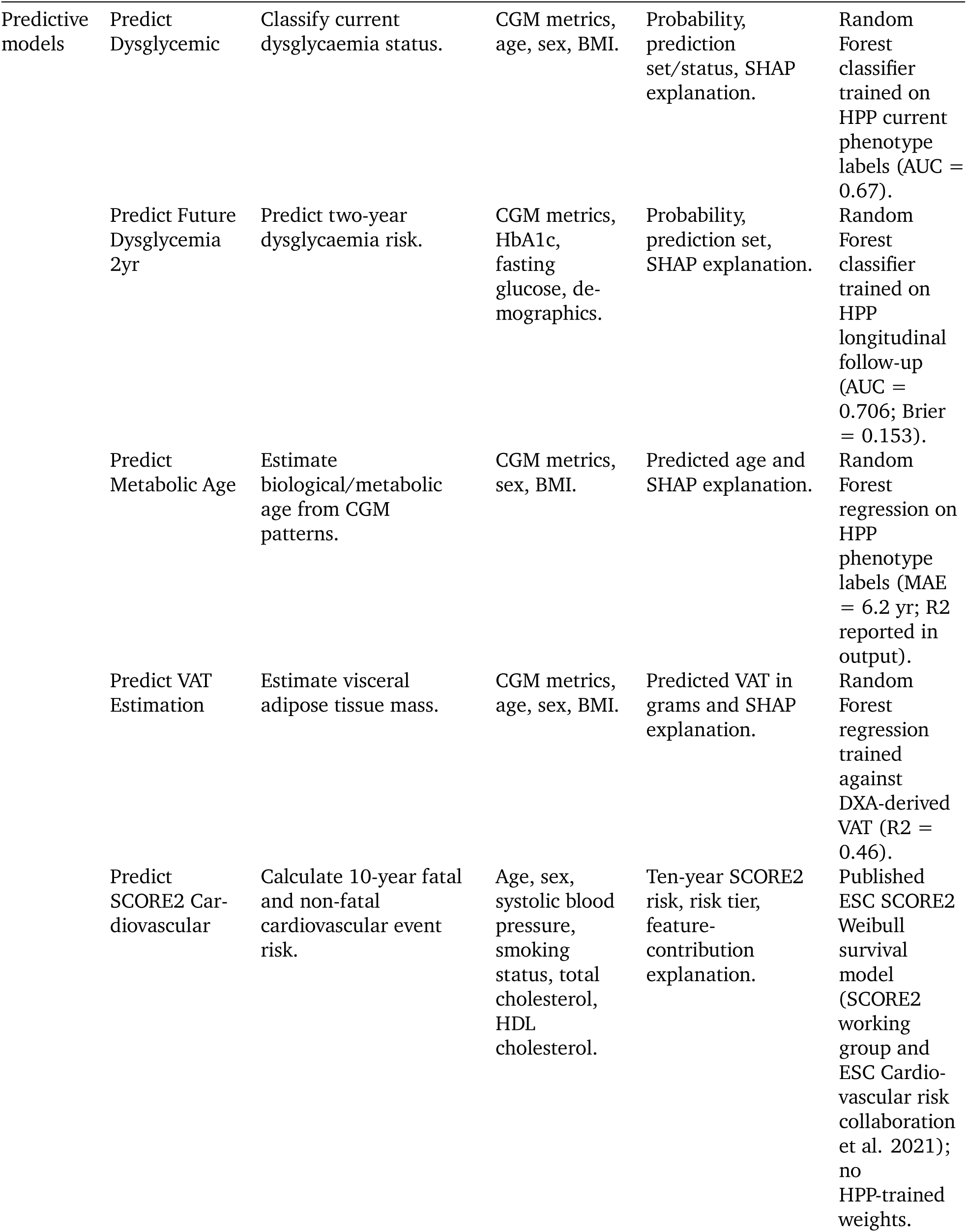
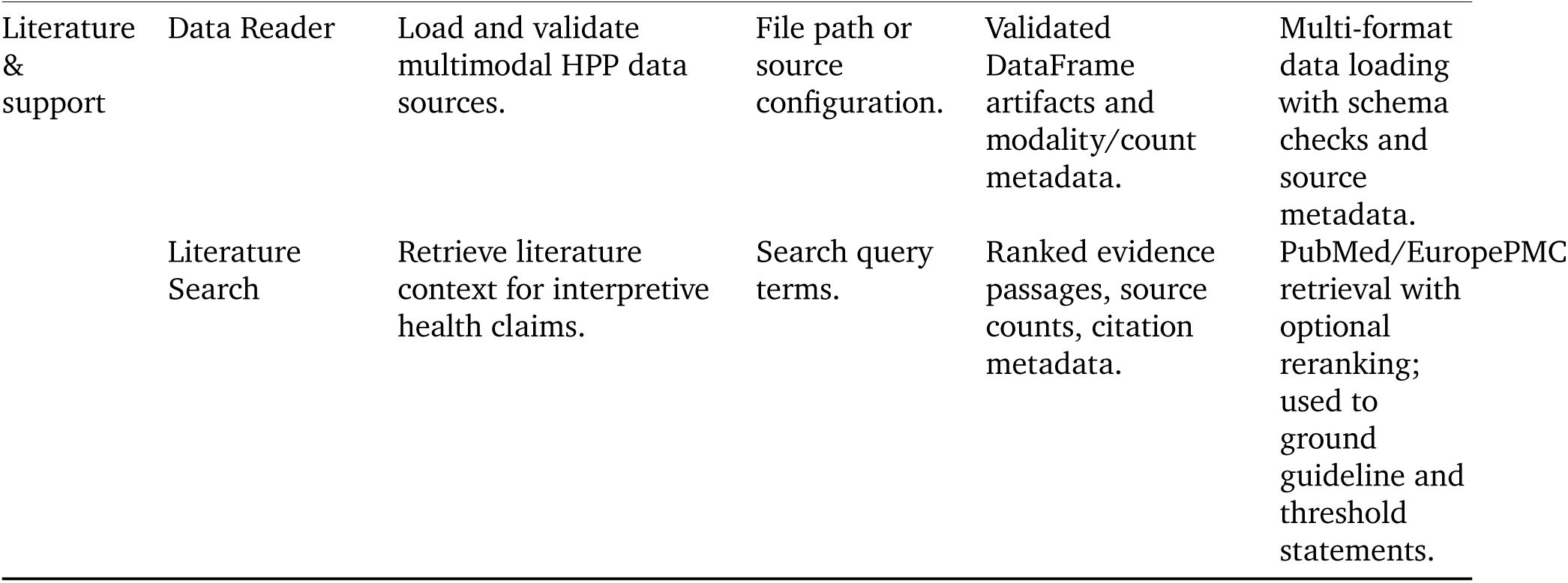

The main experiment disabled fcdb_enrichment and the meal optimiser so that the matrix measured the stable P0 reporting stack rather than optional food-database enrichment or meal-rewriting extensions. These tools remain part of the agent surface because they are relevant for product workflows, but their exclusion is fixed in the experiment contract.

#### S1a. Predictive model details

The agent integrates five predictive models, four trained on HPP data and one implementing a published clinical algorithm.

| Model | Task | Method | Training N | Key Metric |
| --- | --- | --- | --- | --- |
| Dysglycaemia classifier | Current glycaemic status | Random Forest | 23,444 | AUC 0.67 |
| 2-year dysglycaemia risk | Prospective pre-diabetes/diabetes onset | Random Forest | 800 (143 test) | AUC 0.71 |
| Metabolic age | Biological age from CGM patterns | Random Forest | 23,442 (4,186 test) | MAE 6.2 yr (R2 0.09) |
| VAT estimation | Visceral adipose tissue mass | Random Forest | 12,406 (2,250 test) | R2 0.46 |
| SCORE2 cardiovascular risk | 10-year CVD event probability | Weibull (published) | 677,684 (ext.) | C-statistic 0.73 |

All Random Forest models use SHAP for post-hoc explainability, providing feature-level attributions that the agent reports alongside each prediction, such as whether fasting glucose or age were the primary drivers of a risk estimate. The 2-year dysglycaemia model further employs conformal prediction (MAPIE) to produce calibrated 90% prediction sets, giving the agent a principled basis for communicating uncertainty. The SCORE2 tool demonstrates the system’s domain portability: extending from metabolic to cardiovascular risk required only a new tool implementation and skill, with no changes to the orchestration architecture, evaluation framework, or existing tools. The suite also integrates GluFormer (Lutsker et al. 2026), a transformer-based glucose forecasting model trained on HPP CGM data, for short-horizon predictions from CGM history and dietary input, and a postprandial glucose response (PPGR) predictor for meal optimisation. Unlike the classification models above, these operate on raw time-series data rather than pre-computed features.

#### S1b. Representative tool methods

The SCORE2 cardiovascular risk tool (predict_score2_cardiovascular) exemplifies a complementary approach to the machine-learning-based predictive models in the suite. Rather than training a model on HPP data, it implements the published ESC SCORE2 algorithm (SCORE2 working group and ESC Cardiovascular risk collaboration et al. 2021), a Weibull survival model with sex-specific coefficients for estimating 10-year fatal and non-fatal cardiovascular event risk. The tool accepts age, sex, systolic blood pressure, smoking status, total cholesterol, and HDL cholesterol, applies population-mean imputation for missing lipid values, and computes a linear predictor from sex-specific coefficients, including age-interaction terms for cholesterol, blood pressure, smoking, and diabetes. The resulting risk estimate is calibrated to the participant’s geographic region using published recalibration parameters covering four European risk tiers. This tool was included alongside the trained models for two reasons. First, it demonstrates the system’s domain portability: extending from metabolic health to cardiovascular risk assessment required only a new tool configuration and skill. Second, it illustrates the complementarity of formula-based and data-driven approaches within the same standardised interface. The SCORE2 tool uses the identical @tool decorator, returns the same structured envelope, and provides feature-contribution explanations analogous to, though distinct from, SHAP values (Lundberg and Lee 2017), allowing the LLM orchestrator to reason over cardiovascular risk using the same patterns it applies to the ML-based predictions. The algorithm was validated on a development cohort of 677,684 individuals across 45 European cohorts, achieving a C-statistic of 0.73, comparable to or exceeding empirical ML models trained on smaller single-cohort datasets (SCORE2 working group and ESC Cardiovascular risk collaboration et al. 2021).

The population percentile tool (population_percentile) provides age– and sex-adjusted percentile rankings for 46 CGM metrics, the subset of the metrics computed by cgm_metrics for which HPP reference distributions exist, against the HPP cohort (N = 9,405). The reference population is stratified into 16 strata: eight 5-year age bins from 30-35 through 65-70 crossed by two sexes. Percentile reference values (5th, 10th, 25th, 50th, 75th, 90th, 95th) are pre-computed empirically for each metric-stratum combination. For a given participant, the tool identifies the matching age-sex stratum and estimates the percentile by linear interpolation between adjacent quantile-table reference points; values below the 5th or above the 95th percentile are censored to 0 and 100, respectively. The tables store these seven empirical quantiles per metric-stratum rather than the full distribution, so the reported value is a quantile-table interpolation with tail censoring, not an exact empirical rank. The 46 metrics span summary statistics (mean, median, standard deviation), threshold-based measures (time in range 70-180 mg/dL, time above 140/180/250 mg/dL, time below 54/70 mg/dL), glycaemic variability indices (CV, MAGE, MODD, CONGA, ADRR), and composite risk scores (GMI, GRADE, J-index, HBGI, LBGI). Sample sizes per stratum vary substantially, from approximately 10 participants in the youngest bins (ages 30-35) to over 1,100 in the most populated bins (ages 45-50). The tool returns the exact stratum N alongside each percentile value, enabling downstream consumers, including the LLM orchestrator and final report, to caveat comparisons drawn from small reference groups. This transparency is enforced at the evaluation layer: the groundedness evals require that every population comparison disclose the matched cohort size.

The 2-year dysglycaemia risk tool (predict_future_dysglycemia_2yr) estimates the probability that a currently normoglycaemic individual will develop pre-diabetes or diabetes within 24 months. The outcome for both dysglycaemia models is derived from the HPP curated diabetes phenotype, pre-diabetes or diabetes versus normal, assessed at the same visit for the cross-sectional classifier and at the 2-year follow-up visit for this prospective model. Unlike the cross-sectional dysglycaemia classifier (predict_dysglycemic), which diagnoses current glycaemic status from CGM patterns alone (N = 23,444; AUC = 0.67), this model incorporates prospective follow-up outcomes and augments CGM metrics with laboratory values. The feature set comprises 15 variables: 10 CGM metrics (mean glucose, CV, MAGE, GMI, time in range 70-180, time above 180, time below 70, HBGI, LBGI, GRADE), two blood test results (HbA1c and fasting glucose, median-imputed when unavailable), and three demographic variables (age, sex, BMI). The model is a Random Forest classifier (150 trees, max depth 15) with hyperparameters optimised via Optuna (50 trials, TPE sampler, F1 objective) (Akiba et al. 2019). Training used 800 participants from the HPP longitudinal follow-up cohort with confirmed 2-year glycaemic outcomes; 143 held-out participants served as the test set (participant-stratified split, ensuring no individual appears in both sets). On the test set the model achieved an AUC of 0.706 (Brier score 0.153). SHAP analysis (Lundberg and Lee 2017) revealed that fasting glucose (mean |SHAP| = 0.064) and age (0.040) dominate feature importance, followed by MAGE (0.027) and HbA1c (0.026), highlighting the complementarity of CGM-derived variability metrics and traditional laboratory markers for prospective risk stratification. Prediction uncertainty is quantified via conformal prediction (MAPIE SplitConformalClassifier) (Taquet et al. 2022), which produces calibrated 90% prediction sets with guaranteed statistical coverage, providing the LLM orchestrator with a principled basis for hedging risk communications in the final report.

### S2. Representative behavioural skill

The meal_cgm_response skill is reproduced below because it shows how behavioural constraints are specified without changing tool code. The skill names the tools the orchestrator should call, identifies the metadata fields that the analyzer may use, and prohibits common hallucinations. The most important line is the instruction not to use item counts as meal counts; that sentence was added after an observed failure in which a model reported parsed food items as meals.

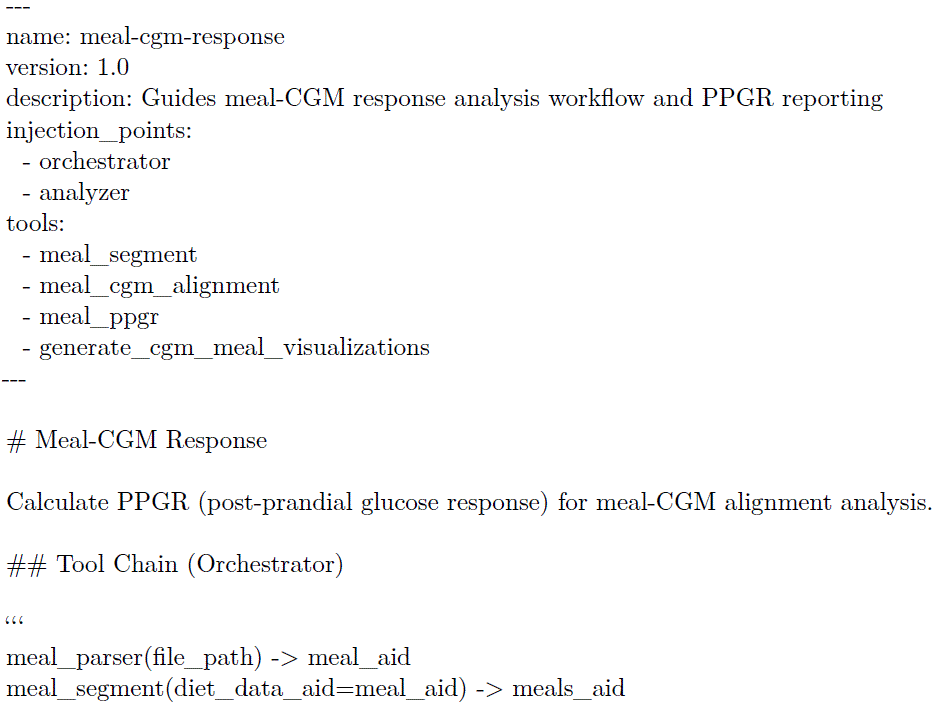

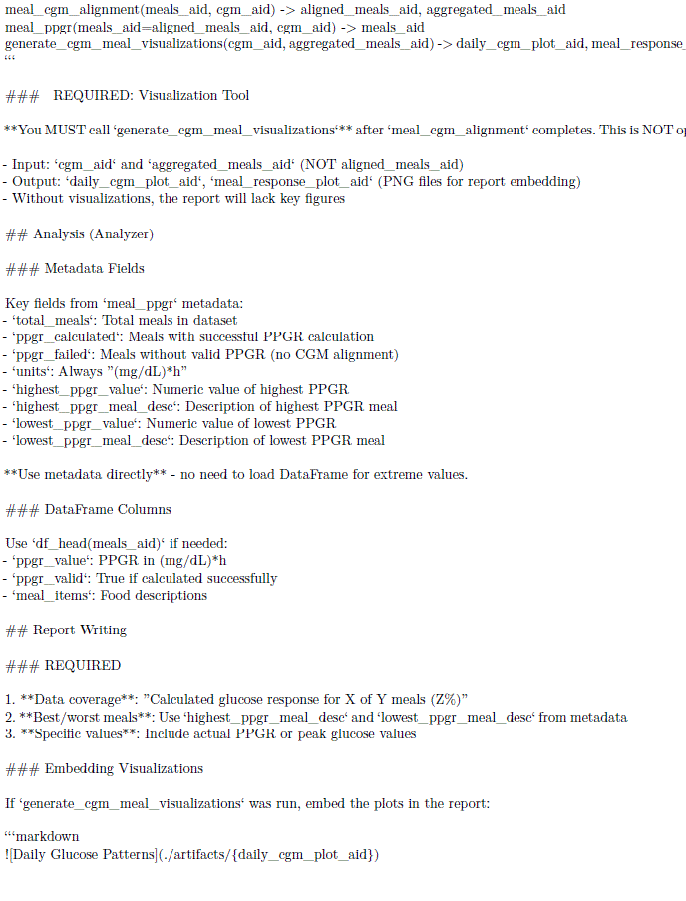

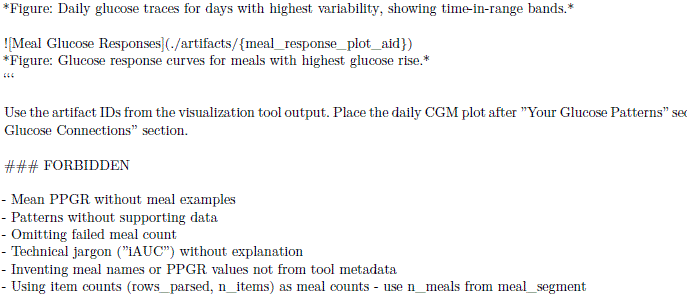

### S3. Eval harness detail

Before writing evals, we specified the tasks they would measure against. For each proposed capability we defined the expected input modalities, required tool chains, and success criteria before writing any agent code. Tasks were stratified at three difficulty levels: easy (computing standard CGM metrics), medium (contextualising a metric with population percentiles and guideline thresholds), and hard (synthesising meal-glucose response patterns alongside longitudinal risk context). Every task bundle included explicit negative tests where the agent should decline, caveat, or acknowledge insufficient data. Selection was biased toward capabilities where HPP cohort data provides a genuine advantage over general-purpose LLMs: population PPGR distributions used for predictor training, matched-cohort comparisons, and predictions from HPP-trained models.

The eval suite grew through a test-driven development cycle rather than being written only after the system was complete. A domain expert (physician and research scientist with dual competence in clinical interpretation and system-level debugging) reviewed generated reports alongside complete tool-calling traces using a purpose-built annotation tool, identifying failures across five dimensions: factual errors, ungrounded claims, clinical language violations, missing caveats, and structural issues. The dual competence mattered: some failures are clinically incorrect, while others are technically correct but misleading. These failure categories emerged empirically rather than from a taxonomy imposed a priori, and directly informed the 21 automated evals.

The report, participant dataset, tool harness, and scoring rules together form an evaluation environment: structurally the same artifact as a reinforcement-learning environment, with a dataset, a harness, and scoring rules (Prime Intellect; Fireworks Eval Protocol). We use it here to measure a fixed policy; the same infrastructure could optimise one. This hierarchy partially maps to granularity levels in related eval work: (Nath et al. 2025) advocates step-wise evaluation of tool-calling sequences, asking whether the agent called the right tools in the right order, corresponding to our deterministic trace checks; (Jiang et al. 2025) achieves programmatic verification in healthcare by scoring agent actions against a structured FHIR environment rather than judging clinical reasoning directly, which our ground-truth comparison evals adopt in spirit. Benchmarks targeting factual medical knowledge (Singhal et al. 2023) or isolated CGM-meal tasks (Das et al. 2025) do not measure whether an agent’s integrated output is coherent, clinically appropriate, and personalised, the gap our framework targets for CGM-based metabolic health.

The eval harness scores every generated report against 21 automated checks. Eighteen are deterministic or ground-truth checks that can run with ––no-llm; three are LLM judges that are reported separately and excluded from the headline form/provenance score. The Figure 4C content/tool-execution split is computed over the five evals configured for that panel: eval_literature_context, eval_digital_twin, eval_meal_ppgr, eval_meal_data_enrichment, and eval_dietary_pattern_completeness. Other deterministic evals also inspect traces or tool meta-data when relevant, including eval_prediction_tool and eval_diet_qc_severity, but they are reported within their primary categories rather than in the Figure 4C split. By contrast, eval_groundedness_llm is narrower: it asks whether glucose event claims such as spikes, lows, or per-day metrics are tied to explicit dates or day references. Baselines are not penalised for failing to execute unavailable tools, but they can still fail content, numerical, language, and citation checks.

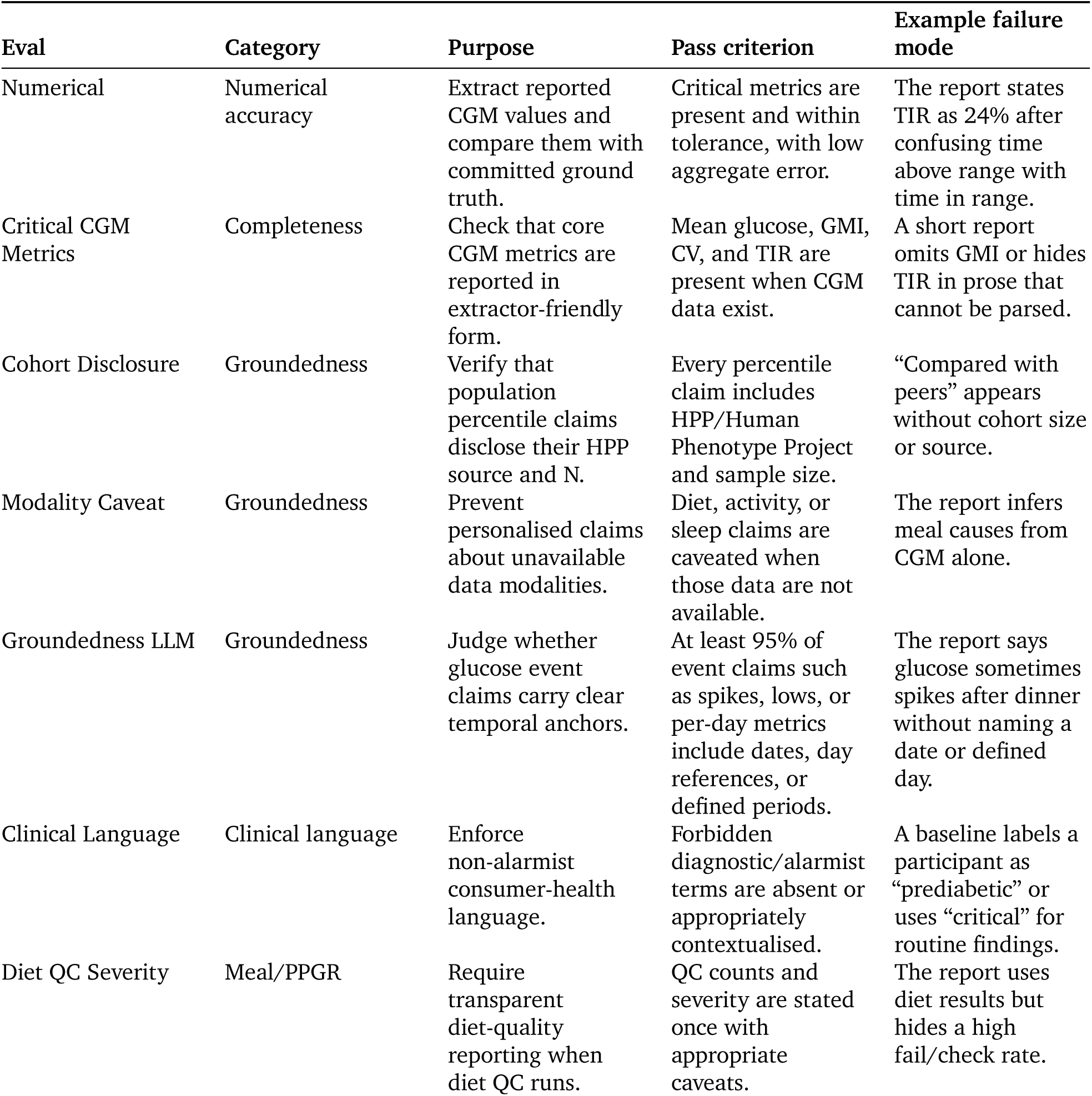
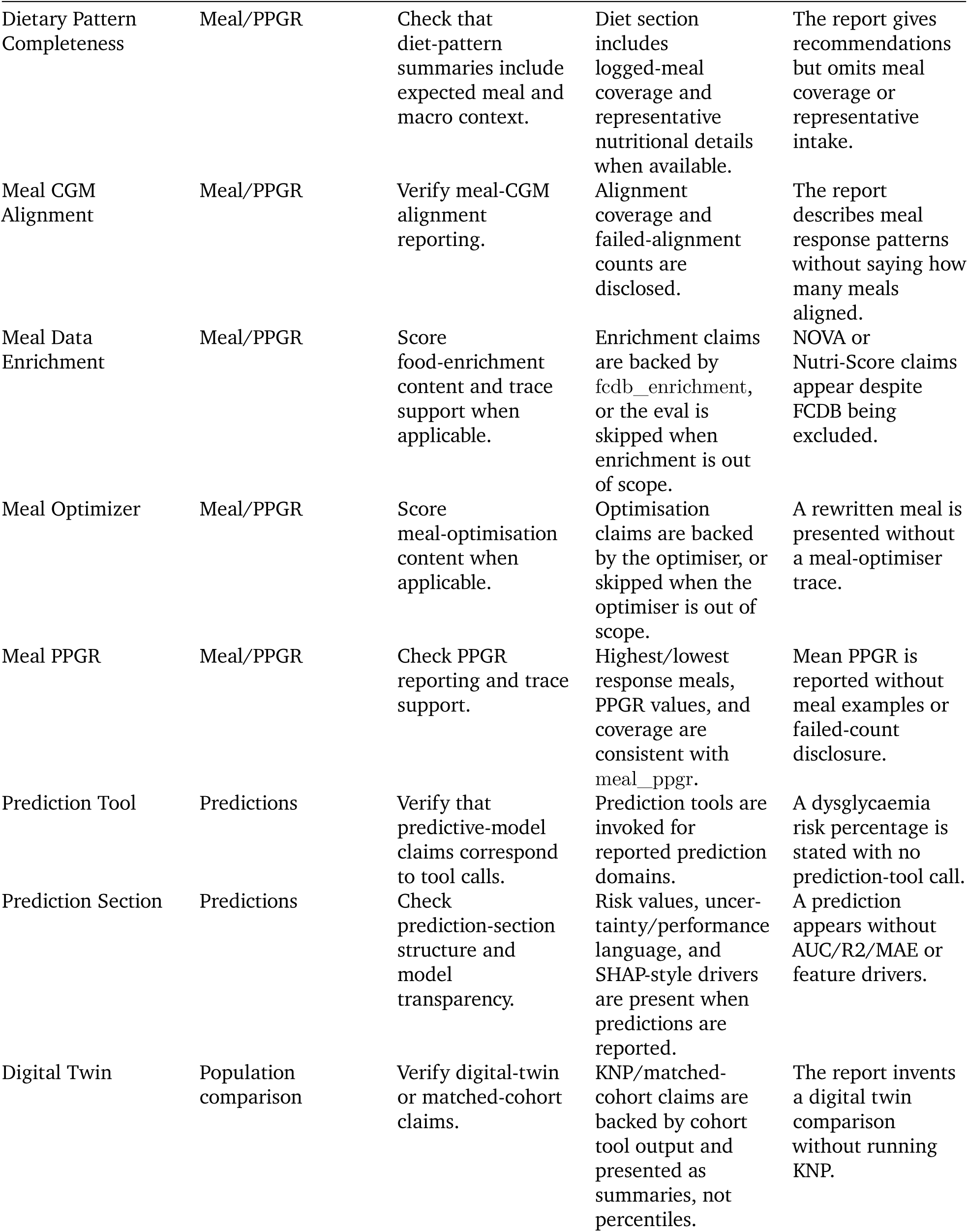
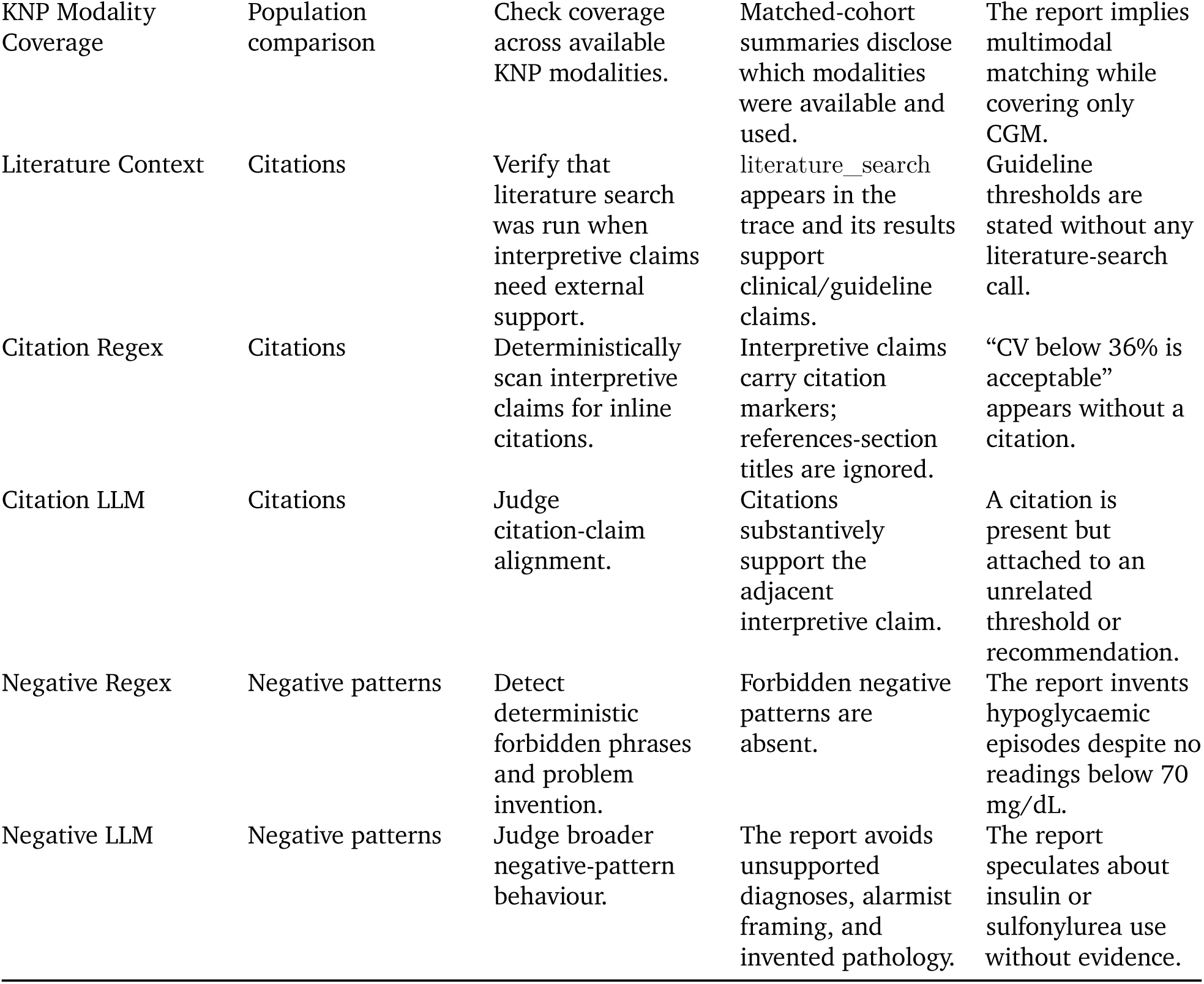

The three LLM judges are retained as supplementary evidence because they behaved differently from the deterministic checks. The citation judge separated conditions cleanly and skill-dependently, whereas groundedness and negative-pattern judges rated even the tool-free baseline highly. This leniency is why the LLM judges are excluded from the headline form/provenance scores and treated as provisional pending judge-versus-human agreement checks.

**Table S3a.**
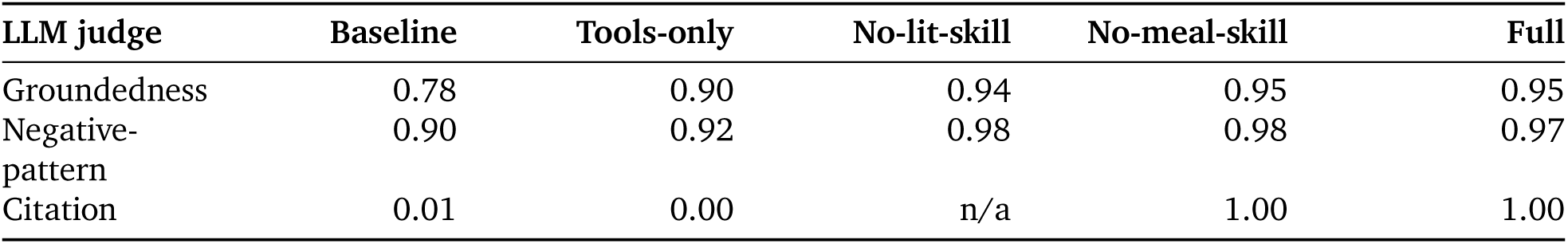
LLM-judge scores by condition (provisional). Mean score for the three LLM judges, pooled across the three prompts (n = 42 reports per condition, except citation is not applicable for the no-lit condition).

### S4. Generalisation Figure

**Figure S2.**
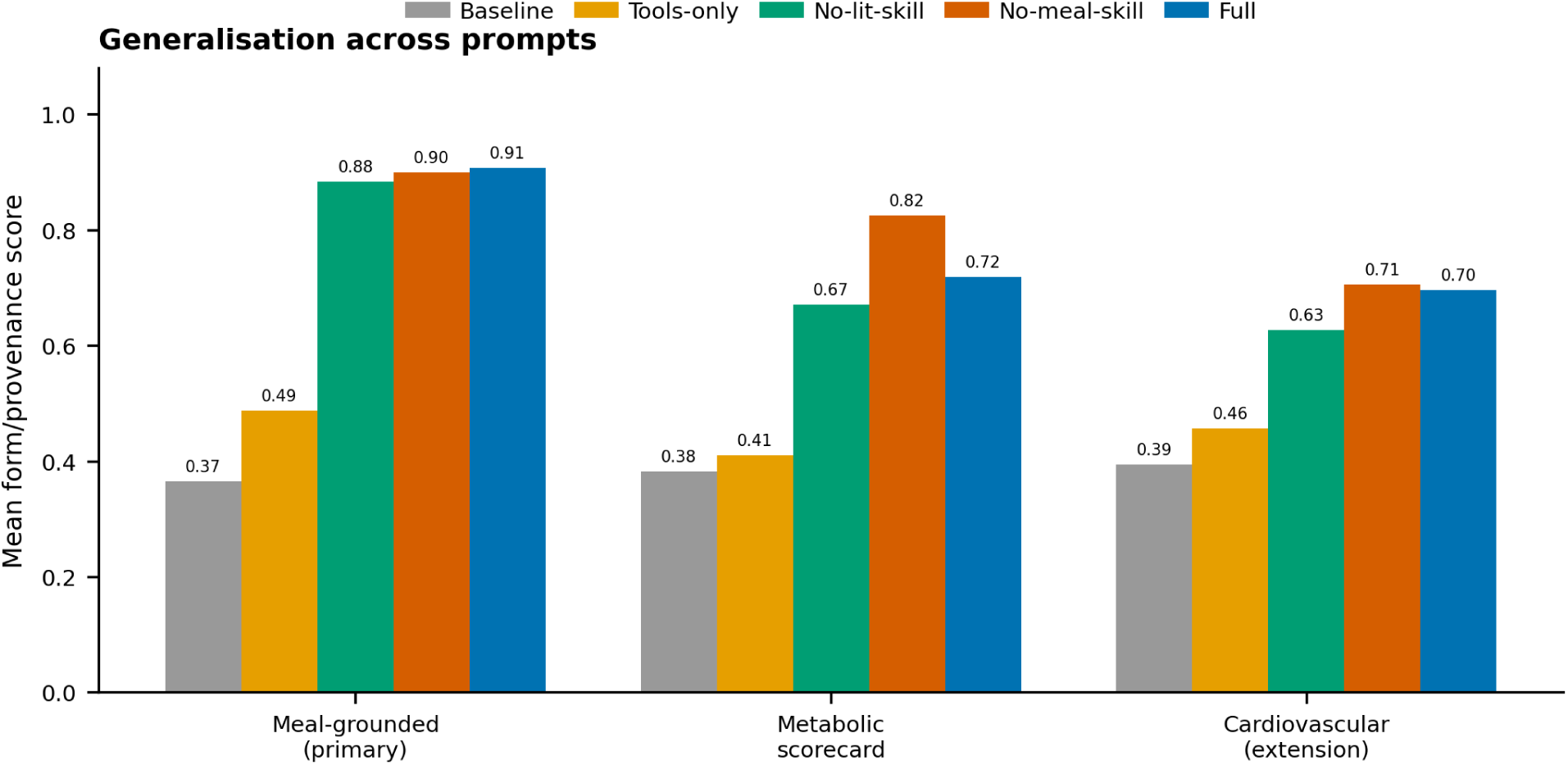
| Generalisation across prompts. Mean form/provenance score for all five conditions on each of the three prompts. The baseline → Full lift is present on all three (Full 0.91 meal-grounded, 0.72 metabolic scorecard, 0.70 cardiovascular), strongest on the primary use case. No-meal-skill exceeds Full on the metabolic scorecard because it is scored over fewer (meal-specific) evals, not because it is better.

### S5. Baseline reproducibility — Participant A (§6.3)

This supplement documents the generation setup for the §6.3 illustrative case study, providing the prompts, model identifiers, provider routes, and settings needed to reproduce or inspect the comparison. The case study is not part of the systematic 14-sample matrix (§6.1–6.2); it examines a single participant (Participant A) across two prompts.

#### S5.1 Participant

Participant A: male, age [X], BMI [X]. Blood markers: fasting glucose [X] mg/dL, HbA1c [X]%. CGM: 14 days (1,248 readings), 100% coverage. Diet logs available. Blood pressure and lipid panel not available at the time of assessment (reason SCORE2 was not computed).

Key validated values (from ground truth and tool outputs):

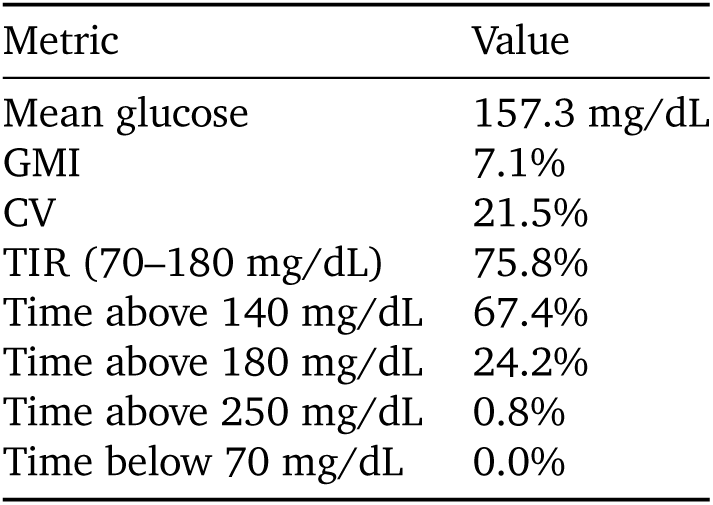

#### S5.2 Prompts

Two prompts were used. The external benchmark models received demographics, blood markers, and CGM time-series as context (no diet logs; diet data is HPP-proprietary and was not included in the benchmark input bundle). The PHA system loaded participant data directly from HPP storage and had full access to CGM and diet logs.

##### Prompt A — metabolic scorecard

Based on my data, give me a quick metabolic scorecard: grade each key area (glucose health, metabolic risk, cardiovascular risk), tell me exactly where I stand compared to people my age and sex, and give me the 2 most important things to change. Keep it to half a page — I want signal, not a report.

##### Prompt B — meal-grounded action

Using my glucose sensor data and diet logs, tell me: which meals are hurting me most, where my metabolic markers put me relative to my peers, and what my cardiovascular risk is. Short answer — just the actionable stuff.

#### S5.3 PHA generation

The PHA reports were generated using the full P0 system (all 21 tools, all 12 skills, Opus 4.6 foundation model).

| Setting | Value |
| --- | --- |
| Foundation model | Claude Opus 4.6 |
| Provider | Amazon Bedrock |
| System | full tools and skills |
| Temperature | Default (Bedrock API default) |
| Seed | Not set |

#### S5.4 External-model generation

External models were queried via OpenRouter with a single-turn user message containing the data bundle (demographics, blood markers, CGM time-series) followed by the prompt. No system prompt was provided. Default sampling settings were used.

| Model label (§6.3) | OpenRouter slug | Temperature | Seed |
| --- | --- | --- | --- |
| GPT-5.4 | openai/gpt-5.4 | default | none |
| Claude Opus 4.6 | anthropic/claude-opus-4.6 | default | none |
| Gemini 3.1 Pro | google/gemini-3.1-pro-preview | default | none |
| Perplexity Sonar Pro | perplexity/sonar-pro | default | none |

Timeout: 180 seconds per call. Requests sent in parallel.

#### S5.5 PHA report for Prompt A

The excerpt below is the Prompt A (metabolic scorecard) output from the full PHA system, corresponding to the “PHA” column of Table 2 in §6.3. Generated with Claude Opus 4.6 on Amazon Bedrock. Some values redacted for participant privacy; all other values are as generated.

##### Interpretation

**Metabolic Scorecard** | M, age [X], BMI [X]

| Domain | Grade | Your number | Peer context |
| --- | --- | --- | --- |
| Glucose control | **F** | Mean 157 mg/dL · GMI 7.1% · TIR 76% | Higher mean glucose than 100% of age- and sex-matched peers (N=298); TIR at 0th percentile |
| Metabolic risk | **C** | Met. age 56.6 yr · Dysglycemia prob. 56% | [X] yr “younger” per model, but current dysglycemia screen positive (56%) offsets that; VAT est. 1546 g |
| Cardiovascular risk | — | Not assessed | BP data unavailable; SCORE2 could not be computed |
| Future trajectory | **F** | 2-yr dysglycemia risk 71% | Top drivers: fasting glucose [X] mg/dL, MAGE 85 mg/dL, HbA1c [X]%, age [X] |

Your CGM pattern is consistent with established type 2 diabetes — mean glucose, variability, and time above 180 mg/dL all exceed every peer in the age-matched male cohort (N = 298). The 2-year model flags a 71% probability of persistent dysglycemia, driven primarily by your fasting glucose ([X] mg/dL) and HbA1c ([X]%). On the positive side, glucose variability (CV 21%) is moderate, and zero time was spent below 70 mg/dL — hypoglycemia risk is currently low.

Meal analysis pinpointed your largest post-meal glucose spike: **Rice + Schnitzel + Vegetable pie + Salad** on [date] at dinner — PPGR 161.1 (mg/dL)·h. By contrast, a lunch of Spaghetti Bolognese + Wholemeal Crackers + Pastrami produced a PPGR of only 0.45, roughly 350× lower.

##### Recommendations

**2 priorities**

1. **Replace white rice at dinner with a lower-glycemic starch.** Your top spike meal centered on rice. In T2D patients, substituting fiber-fortified rice or reduced-carbohydrate noodles lowered 2-h iAUC from 160 to 114 mmol/L·min (−29%, p = 0.046; Yusri et al. 2025, doi:10.3389/fnut.2025.1594890). Eating vegetables and protein before the starchy portion further blunts the response (Papakonstantinou et al. 2022, doi:10.3390/nu14040823).
2. **Add 150 min/week moderate-intensity activity** to target fasting glucose — the single largest contributor to your 2-year risk. Even modest weight loss meaningfully improves insulin sensitivity in this age group.

**Not assessed:** ApoB, Lp(a), blood pressure, lipid panel, family history, renal function — cardiovascular risk grading requires these.

**Confidence:** Glucose control HIGH (13 days CGM, 0 gaps, N=298 reference) | Metabolic risk LOW (metabolic age R²=0.09; dysglycemia AUC=0.67) | Future trajectory MEDIUM (AUC=0.71, lab-confirmed HbA1c available)

### S6. Representative cardiovascular risk report (Participant B)

The report below is a representative cardiovascular risk assessment generated by the system for Participant B, with the sample identifier anonymised. All metric values are as generated; the narrative rounds tool outputs — precise tool-trace values: metabolic age 50.30 yr (narrative: “50 years”), 2-year dysglycaemia risk 24%, CVD risk 3.56% (narrative: “3.6%”). Some values redacted for participant privacy.

## Cardiovascular Health Assessment

### Executive Summary

**Cardiovascular-Metabolic Health**: A (High Confidence)

Your comprehensive cardiovascular profile is excellent: glucose stability in the top 5% of peers, metabolic age [X] years younger than chronological age, and low 10-year cardiovascular disease risk (3.6%). All assessed domains show favorable patterns.

**Assessed:** Glycemic health (13-day CGM + HbA1c), metabolic age, dysglycemia trajectory, 10-year CVD risk (SCORE2 with complete data including cholesterol)

**Not assessed:** apoB, Lipoprotein(a), coronary artery calcium score, family history

**Key action:** Consider one-time Lp(a) test to assess genetic cardiovascular risk

### Data Scope

This assessment evaluates cardiovascular-relevant metabolic health based on:

- **Continuous glucose monitoring** (13 days, 1,248 readings)
- **Blood biomarkers:** HbA1c [X]%, fasting glucose [X] mg/dL, total cholesterol [X] mg/dL, HDL [X] mg/dL
- **Clinical measurements:** Blood pressure [X] mmHg (systolic), BMI [X], non-smoker
- **Population comparison:** Age/sex-matched reference (N=616 males, age [X])

**Limitations:** Does not assess advanced lipid markers (apoB, Lp(a)), coronary calcium, or family history. These factors can independently influence cardiovascular risk and should be considered in comprehensive clinical evaluation.

### Component Scores

| Component | Grade | Data Source | Confidence |
| --- | --- | --- | --- |
| Glycemic Health | **A** | CGM (13 days) + HbA1c | High |
| Metabolic Health | **B+** | Predictive model<br>($R^2=0.09$) | Medium |
| Future Risk Trajectory | **A** | Predictive model<br>(AUC=0.71),<br>age-adjusted | Medium |
| CV Risk (SCORE2) | **A** | Clinical formula<br>(complete data) | High |
| **Overall CV Health** | **A** | **Complete assessment** | **High** |

***Glycemic Health: A (High Confidence)***

***Your glucose control is exceptional — better than 94.5% of age-matched males***.

- **HbA1c:** [X]% (optimal range)
- **Glucose variability (CV):** 10.85%, placing you at the 5.5th percentile among 616 age-matched males
- **Time in range (70-180 mg/dL):** 99.84% — virtually perfect
- **Mean glucose:** 97.76 mg/dL (51st percentile, healthy mid-range)
- **Glucose management indicator (GMI):** 5.65% (52nd percentile)

**What this means:** Low glucose variability is a strong protective factor for cardiovascular health. Your stable glucose patterns reduce oxidative stress and inflammation, key drivers of atherosclerosis. The combination of low HbA1c and exceptional stability is rare in this age group.

**Key protective factor:** Your MAGE (mean amplitude of glycemic excursions) of 28.5 mg/dL is well-controlled, contributing –2.05 years to your metabolic age calculation.

***Metabolic Health: B+ (Medium Confidence)***

***Your metabolic age is 50 years — younger than your chronological age ([X])***.

- **Predicted metabolic age:** 50.3 years (model R²=0.09, indicating high prediction uncertainty but favorable point estimate)
- **Estimated visceral adipose tissue (VAT):** 834.8 cm³ (moderate for age/sex, driven primarily by age rather than adverse metabolic patterns)
- **Current dysglycemia risk:** 7% — very low (AUC=0.67)
- **BMI:** [X] (normal weight, upper range)

**What this means:** While the metabolic age model has low overall explanatory power (R²=0.09), your point estimate of 50 years is favorable. Your glucose stability (MAGE, CV) and healthy BMI are the primary contributors to this younger metabolic age. The VAT estimation is typical for your age and sex.

**Top contributors to favorable metabolic age**:

1. **MAGE** (28.5 mg/dL): Contributes –2.05 years
2. **BMI** ([X]): Contributes –0.16 years
3. **Low glucose variability (CV):** Minimal adverse contribution (+0.30 years)

**Note on confidence:** The wide model uncertainty (R²=0.09) means metabolic age should be interpreted as a general indicator rather than a precise measure. Your favorable glucose patterns provide more direct evidence of good metabolic health.

***Future Risk Trajectory: A (Medium Confidence, Age-Adjusted)***

**Your 2-year dysglycemia risk of 24% is LOW for males of similar age**.

- **2-year dysglycemia probability:** 24% (model AUC=0.71)
- **Age-adjusted interpretation:** This risk is well below typical for your age group. While absolute risk increases with age (non-modifiable), your modifiable factors (glucose control, BMI, current HbA1c) are all favorable.

**What this means:** Age is the strongest predictor of future dysglycemia (+5% contribution), but your protective factors substantially offset this:

- **Fasting glucose** [X] mg/dL: Contributes –4% (protective)
- **HbA1c** [X]%: Contributes +2% (minimal risk)
- **Glucose variability (MAGE):** Contributes –1% (protective)

**Key insight:** A 24% 2-year risk may sound concerning in isolation, but it reflects normal aging biology. Compared to age-matched peers, you are in a favorable position. Maintaining current patterns will help preserve this advantage.

***Cardiovascular Risk (SCORE2): A (High Confidence)***

**Your 10-year cardiovascular disease risk is 3.6% — LOW RISK according to European Society of Cardiology guidelines**.

- **SCORE2 10-year CVD risk:** 3.56% (validated clinical formula, not ML model)
- **Risk category:** Low risk (<5%)
- **Complete data available:** Age, sex, blood pressure, cholesterol (total and HDL), smoking status

**What this means:** SCORE2 estimates your probability of a major cardiovascular event (heart attack, stroke) over the next 10 years. At 3.6%, you are in the low-risk category, which is excellent for a male of similar age.

**Top protective factors**:

4. **Non-HDL cholesterol:** [X] mg/dL — favorable, contributes –1.11% to risk
5. **HDL cholesterol:** [X] mg/dL — protective, contributes –1.04% to risk
6. **Non-smoker status:** Major protective factor
7. **Blood pressure:** [X] mmHg systolic — slightly elevated but contributes minimally (+1.02%)

**Age and sex contributions:** At your age and being male inherently contributes ∼6% to baseline risk (age +4%, male sex +2%), but your favorable modifiable factors bring total risk down to 3.6%.

**Clinical context:** According to ESC 2021 guidelines, low-risk individuals (<5%) should focus on maintaining healthy lifestyle patterns. Preventive medication is generally not indicated at this risk level unless other high-risk features are present.

## Longevity Outlook

### Your cardiovascular-metabolic profile suggests a highly favorable trajectory for long-term health

Your exceptional glucose stability (top 5% of peers) is your strongest protective asset. Stable glucose reduces cumulative damage to blood vessels, heart, and kidneys over decades. Combined with low 10-year CVD risk, favorable lipid profile, and metabolic age [X] years younger than chronological age, you are well-positioned for healthy aging.

### Key protective patterns to maintain

- Glucose variability control (CV 10.85%, MAGE 28.5 mg/dL)
- Healthy body composition (BMI [X])
- Non-smoking status
- Favorable cholesterol ratio (HDL [X] mg/dL, non-HDL [X] mg/dL)

**Natural aging considerations:** While your 2-year dysglycemia risk (24%) and metabolic age will naturally trend upward with aging, your current metabolic resilience provides a buffer. The goal is to slow the rate of increase, not eliminate age-related changes entirely.

### What This Means for You

#### Strengths to Maintain

8. **Glucose stability** — Your CV of 10.85% and 99.84% time in range are exceptional. Whatever dietary and lifestyle patterns you’re following are working extremely well.
9. **Lipid profile** — HDL of [X] mg/dL and total cholesterol of [X] mg/dL are both favorable. Your non-HDL cholesterol ([X] mg/dL) is well within optimal range.
10. **Metabolic resilience** — Metabolic age [X] years younger than chronological age reflects favorable cumulative metabolic health.
11. **Non-smoking** — This single factor has profound long-term benefits for cardiovascular and overall health.

#### Opportunities for Fine-Tuning

12. **Blood pressure** — At [X] mmHg systolic, you’re in the “elevated” range (120-129 mmHg per ACC/AHA guidelines). While this contributes minimally to your SCORE2 risk, targeting <120 mmHg through lifestyle modifications (sodium reduction, DASH diet, regular aerobic exercise) could provide additional cardiovascular benefit.
13. **Advanced lipid assessment** — Your calculated non-HDL cholesterol ([X] mg/dL) is favorable, but apoB and Lipoprotein(a) testing would provide more precise risk stratification. apoB measures particle number (more predictive than LDL-C in some individuals). Lp(a) is a genetic risk factor independent of lifestyle — one-time test recommended.
14. **Body composition** — BMI [X] is at the upper end of normal range. Depending on muscle mass and fat distribution, there may be room for optimization, though your low VAT estimate and excellent glucose control suggest healthy body composition.

## Recommended Next Steps

***Priority 1: Advanced Lipid Markers (One-Time)***

**Lipoprotein(a) [Lp(a)] testing**

- **Why:** Genetic cardiovascular risk factor, independent of cholesterol and lifestyle
- **Who:** Recommended for all adults at least once (ESC guidelines)
- **Interpretation:** Lp(a) >50 mg/dL (∼30% of population) significantly increases CVD risk
- **Action if elevated:** More aggressive LDL/apoB targets, consider preventive therapies

**Apolipoprotein B (apoB)**

- **Why:** More precise measure of atherogenic particle burden than LDL cholesterol
- **When:** Particularly valuable if triglycerides elevated or metabolic syndrome present (not your case, but completes the picture)
- **Target:** <80 mg/dL for low-risk individuals, <65 mg/dL for moderate risk

***Priority 2: Coronary Artery Calcium (CAC) Score (Optional)***

**Consider if**:

- Family history of premature CVD (age <55 in males, <65 in females)
- Desire for more precise risk stratification
- Borderline treatment decisions

**Why:** CAC score of 0 (absent calcification) is highly reassuring; score >100 may warrant more intensive risk factor management even with low SCORE2.

**Not urgent** given your current low-risk profile, but useful for personalized risk assessment.

***Priority 3: Blood Pressure Optimization***

**Target**: <120/80 mmHg through lifestyle

**Strategies**:

- Reduce sodium intake to <2,300 mg/day (ideally <1,500 mg/day)
- DASH diet (rich in fruits, vegetables, whole grains, low-fat dairy)
- Regular aerobic exercise (150+ min/week moderate intensity)
- Maintain healthy weight
- Limit alcohol consumption

**Monitor:** Home blood pressure monitoring to confirm office readings and track progress

***Priority 4: Continue Current Patterns***

Your glucose control, body composition, and overall metabolic health are exceptional. Whatever dietary and lifestyle habits you currently maintain are highly effective — continue them.

**References and Evidence Base**

**SCORE2 Risk Assessment:**

SCORE2 W-orking Group and ESC Cardiovascular Risk Collaboration (2021). “SCORE2 risk prediction algorithms: new models to estimate 10-year risk of cardiovascular disease in Europe.” European Heart Journal, 42(25), 2439-2454. DOI: 10.1093/eurheartj/ehab309

**Glucose Variability and Cardiovascular Risk:**

Ceriello, A., et al. (2008). “Glycemic variability: both sides of the story.” Diabetes Care, 31(Suppl 2), S272–S278. DOI: 10.2337/dc08-s245

Nalysnyk, L., et al. (2010). “Glycemic variability and cardiovascular disease: a systematic review and meta-analysis.” Diabetes, Obesity and Metabolism, 12(4), 288–298.

**Advanced Lipid Testing:**

Mach, F., et al. (2020). “2019 ESC/EAS Guidelines for the management of dyslipidaemias: lipid modification to reduce cardiovascular risk.” European Heart Journal, 41(1), 111–188. DOI: 10.1093/eurheartj/ehz455.

**Population Comparison:**

Human Phenotype Project database (N=616 age-matched males, glucose variability metrics)

**Quality Assurance Notes**

**Data Confidence Summary**:

- **Glycemic assessment:** HIGH (13 days CGM + HbA1c)
- **Metabolic age:** MEDIUM (model R²=0.09 indicates high uncertainty, but point estimate favorable)
- **Future dysglycemia risk:** MEDIUM (model AUC=0.71, age-adjusted interpretation)
- **SCORE2 CVD risk:** HIGH (complete data, validated clinical formula)
- **Overall assessment confidence**: HIGH

**Model Metrics Reported**:

- Metabolic age: R²=0.09 (low explanatory power, wide prediction intervals expected)
- Current dysglycemia: AUC=0.67 (fair discrimination)
- Future dysglycemia (2-year): AUC=0.71 (good discrimination)
- SCORE2: Formula-based calculation (AUC=0.73 from validation studies), not machine learn-ing

**Limitations Acknowledged**:

- Metabolic age model has low R² — interpret as general indicator
- No assessment of apoB, Lp(a), family history, or coronary calcium
- SCORE2 most validated for ages 40-69 in European populations
- CGM period (13 days) represents snapshot; longer monitoring confirms stability

**No Diagnostic Claims:** This assessment provides risk stratification and health optimization guidance. It does not diagnose disease or replace clinical evaluation.

